# How to measure obesity in public health research? Problems with using BMI for population inference

**DOI:** 10.1101/2025.04.01.25325037

**Authors:** Adam Visokay, Kentaro Hoffman, Stephen Salerno, Tyler H. McCormick, Sasha Johfre

## Abstract

**Background:** Though viewed as problematic for measuring individual-level adiposity, Body Mass Index (BMI) is often considered “good enough” for population inference and epidemiological research. However, we demonstrate that BMI produces statistically invalid population-level estimates of associations between key demographic risk factors (e.g., self-reported sex, race, age) and obesity when compared to more direct adiposity measurements. Further, we demonstrate how novel statistical calibration techniques can enable more valid population inference using widely available BMI data alongside a limited subset of “gold standard” measurements.

**Methods:** Using National Health and Nutrition Examination Survey data (2011-2023), we compare associations, broken down by demographic groups, across three different purported adiposity measures: BMI, Waist Circumference (WC), and whole-body total fat percentage from Dual-energy X-ray absorptiometry (DXA) scans. We then apply a statistical procedure for conducting inference on predicted data to calibrate BMI-based prevalence estimates toward the “gold standard” DXA-based measurements, allowing for valid population inference even for time periods where only BMI data are available.

**Findings:** BMI-measured adiposity yields substantially different – and often contradictory – conclusions about the association between obesity status and lifestyle factors compared to the more direct, DXA-based measurements. Most concerning, the directions and magnitudes of the associations between racial groups may differ depending on whether BMI or DXA-based measurements are used. Similarly, self-reported sex-based differences in obesity prevalence show opposite patterns across measurement types. Our validation results confirm that our calibration method overcomes this challenge and successfully approximates DXA-based associations using primarily BMI-based measurements.

**Interpretation:** Our study provides empirical evidence that uncorrected BMI-based inference leads to invalid population-level estimates about the associations between obesity status and key predictors. The statistical calibration approach we present offers a practical solution for obesity researchers who must rely on BMI or similar anthropometric measures due to cost or data availability constraints, enabling more valid population inference without requiring comprehensive “gold standard” adiposity measurements.

## Introduction

Body Mass Index (BMI; kg/m^2^) is a simple and inexpensive medical screening tool for obesity, derived from a person’s height and weight. The current “gold standard” measure for whole-body fat percentage is a dual-energy X-ray absorptiometry scan (DXA), which provides a precise breakdown of bone density, lean muscle mass and visceral and subcutaneous fat mass, but are either prohibitively expensive or inaccessible for most large-scale population studies.^1–3^ Despite being a crude predictor of adiposity at best, the BMI’s ease of use has led to its dominance in health research, as it is often assumed to measure “fatness” directly.^4, 5^ BMI is also known to be correlated with several morbidities as well as overall and cause-specific mortality – but as many before us have pointed out, this correlation is more apparent for some conditions and in some populations than others.^6–8^

Therefore, it is crucial to recognize that BMI *predicts* adiposity, but does not *directly measure* it. This fundamental distinction is made clear in Rothman et al.’s canonical textbook, “Modern Epidemiology”,^1^ but it tends to be framed as a limitation only in the clinical diagnosis of individuals, not for population-based studies.^9^ However, our findings challenge this prevailing assumption that BMI is an adequate predictor for population-level and epidemiological research. We demonstrate that prediction errors from BMI significantly bias inference at the population level as well, undermining its reliability across both individual and aggregate analyses. Building on this insight, we apply a new paradigm for understanding the role and limitations of BMI in obesity research. We consider BMI to be one of several potential *adiposity prediction algorithms* (see Bergman 2011 and Swainson 2017 for several more examples^10, 11^), where an *algorithm* is simply a set of step-by-step decision rules that map inputs to outputs. In this framework, BMI takes as input height and weight and gives as output a “healthy-weight” category. Figure 1 demonstrates the full flowchart, including all intermediate steps.

**Figure 1:**
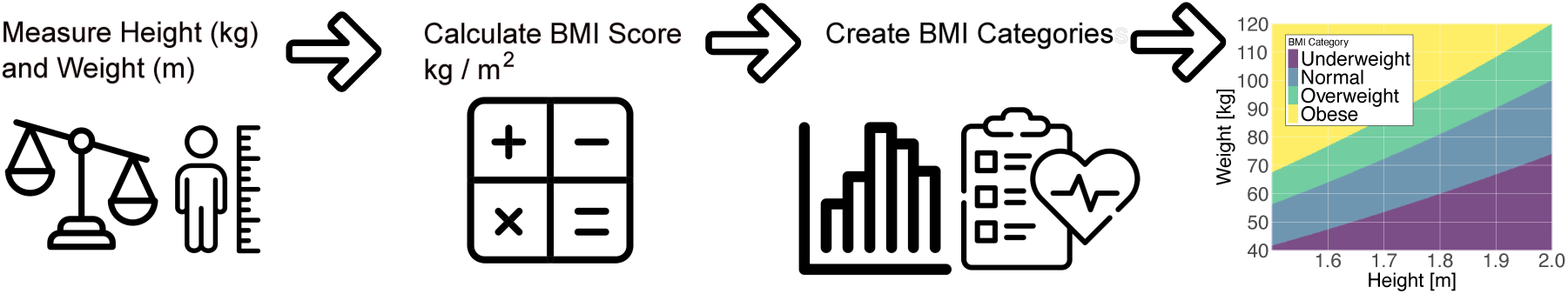
The Body Mass Index is an adiposity prediction algorithm. Continuous measures of height and weight are measured for each individual. Then, a continuous BMI score is calculated. Finally, that continuous score is discretized into categories. Before the 1990’s, the BMI categories were based on statistical distributions of BMI scores (quartiles or quintiles). Over time, the categories were standardized according to associations between BMI score and morbidities like type 2 diabetes.

**Figure 2:**
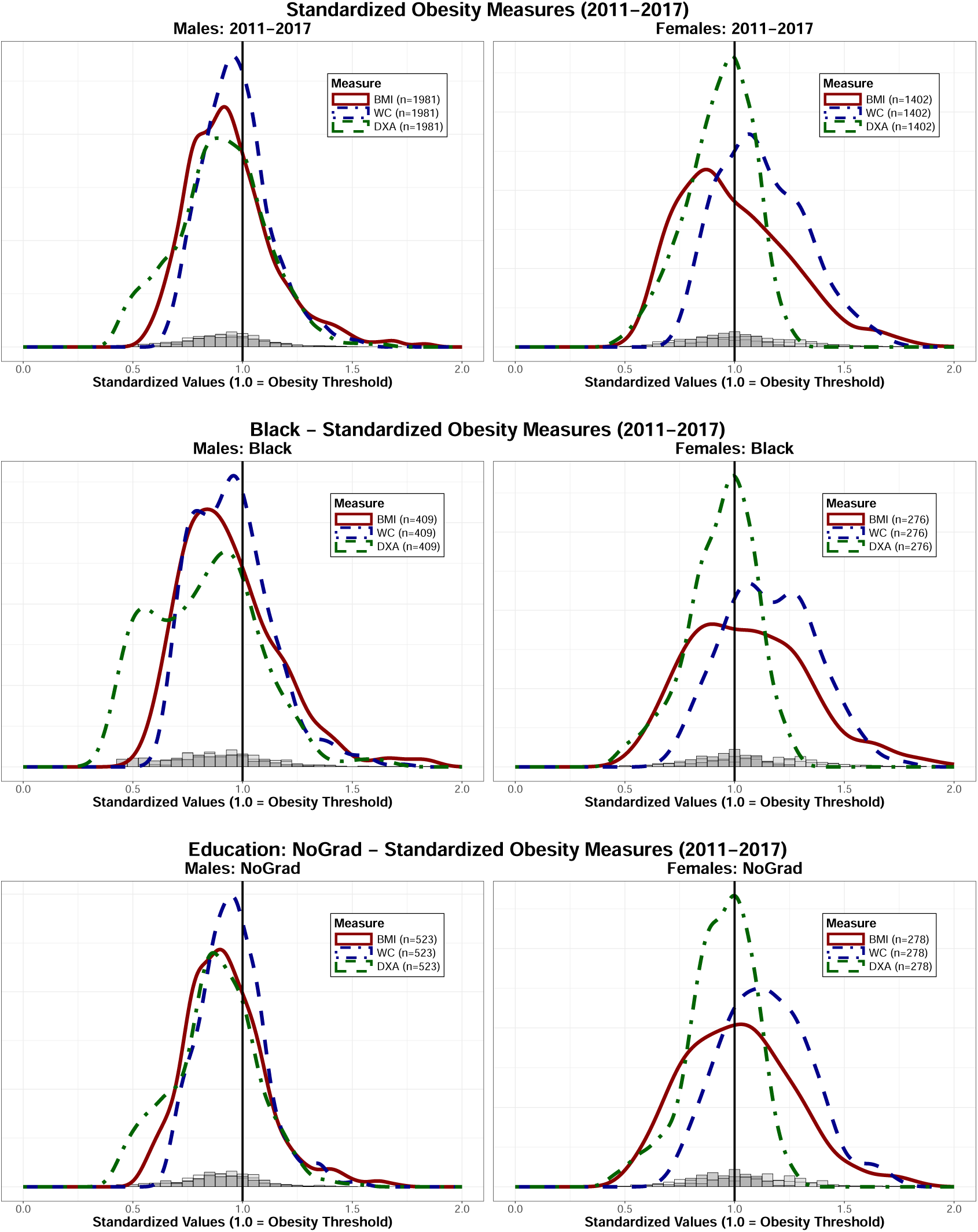
Density plots for the distribution of each continuous measure of adiposity are shown above, decomposed by sex, sex and black race/ethnicity, and sex and less than a high-school education. Individuals with a standardized value greater than 1.0 are considered obese according to the associated measure. BMI and WC are most similar to the “gold standard” DXA measure for males. These anthropometric measures are considerably different from DXA for female respondents, especially black females.

Conceptualizing BMI as a prediction algorithm has two crucial implications. First, recognizing that predictions introduce bias and uncertainty into statistical inference allows us to quantify the bias in associations between risk factors (e.g. age, sex, race) and obesity that may arise solely from the adiposity measure we use (e.g., BMI versus WC versus DXA). These statistical relationships may vary in sign and in magnitude as individuals with different characteristics are misclassified at different rates. Numerous studies have documented how BMI misclassifies muscular individuals as obese and fails to identify metabolically unhealthy “normal-weight” individuals.^12, 13^ While the public health community has increasingly acknowledged these limitations for individual diagnoses, many researchers continue to incorrectly assume that anthropometric measures like BMI or waist circumference adequately represent adiposity for population-level analyses.^9, 14–16^ Second, framed this way, we can draw upon recent advances in performing valid “inference with predicted data” (IPD) for insights into how we can methodologically address this challenge using statistical calibration.^17–24^

The Lancet’s recent Diabetes & Endocrinology Commission on obesity has proposed a more comprehensive definitional framework,^25^ but even improved prediction models cannot resolve the fundamental measurement issue: predictions cannot be used in place of direct measures for population research without an appropriate calibration. In the following analysis, we demonstrate BMI’s poor performance in population inference compared to DXA and highlight the statistical consequences of this misspecification. Then, we present a novel application of IPD correction to directly address this challenge. Our solution enables researchers to conduct valid population-level inference for adiposity-related outcomes while utilizing widely available BMI data.

Here, we examine obesity as a dependent variable and illustrate how obesity classifications based on different adiposity measurements can yield divergent scientific conclusions. We specifically compare how common predictors (self-reported sex, race, age, household income, education, and smoking status) are associated with three supposed adiposity measures: BMI score, waist circumference (WC) and percentage of total body fat measured by DXA. For example, when investigating the research question “How does obesity associate with different racial groups?”, we assess whether the conclusions vary depending on which adiposity measure is used. We then apply an IPD calibration specifically designed to reduce the bias that occurs when using predicted data instead of preferred direct measurements. This provides a practical solution for obesity research contexts where only inexpensive anthropometric measures, such as BMI, are available. While our analysis primarily highlights the contrasts between BMI and DXA measurements, we include WC in our analysis because it has demonstrated superior predictive capacity for some health outcomes compared to BMI.^1, 26^

## Data

We use data from the National Health and Nutrition Examination Survey (NHANES) to measure the association between obesity and common demographic and behavioral predictors.^27^ For several survey cycles, from 2011-2017, NHANES measured whole-body fat percentage with DXA scans as well as BMI and WC. Data collection for NHANES 2021-2023 was interrupted by the COVID-19 pandemic, so for those years, only BMI and WC were collected. We convert BMI scores, WC measurements and DXA measured total body fat percentage to a binary outcome – obese or not – by labeling anyone with BMI*>*30kg/m^2^, males with WC*>*88cm, DXA*>*30% and females with WC*>*102cm and DXA*>*42% as obese and everyone else as not-obese.^28, 29^ One could instead use other thresholds as cutoffs for obesity, such as the original BMI-obesity thresholds adopted in 1998 (27.8 for men and 27.3 women).^30^ Or, as others have suggested, one could use ethnicity-specific BMI categories, or continuous measurements and fit a linear regression. Another option could be to convert the continuous BMI scores into ordinal BMI categories, as shown in Figure 1, instead. We include several of these analyses as robustness checks with additional discussion in the Appendix A.5, A.6. However, none of these considerations change the substantive conclusion: while DXA is a measure of adiposity, BMI and WC are measures of *something else*, and this leads to invalid inference more prominently for some groups than others.

## Methods

Treating algorithmically-derived values as directly measured outcomes can lead to biased results and incorrect substantive conclusions.^17, 31–34^ This is especially concerning when such analyses guide resource allocation or public policy decisions. Several methods for performing inference with predicted data (IPD) have been proposed (see Hoffman et al. 2024 for a brief overview).^19^ Conceptualizing BMI as an obesity prediction algorithm allows us to adopt this IPD framework and develop approaches for more accurate statistical inference about obesity using BMI.

In an ideal research scenario, we would utilize DXA-measured whole-body percentage fat for every individual when studying obesity as a health condition, as it provides the most accurate assessment of adiposity. NHANES data from 2011-2017 include DXA measurements, allowing us to conduct such “gold standard” adiposity analyses for this period. However, DXA data are not available for the 2021-2023 NHANES wave (due to COVID-19 interruptions and resource constraints), creating a critical gap in our ability to perform accurate inference for recent populations.

This data availability challenge exemplifies the broader practical constraints researchers face – DXA scans are prohibitively expensive or inaccessible for many large-scale studies, forcing reliance on more economical alternatives like BMI. Our analysis demonstrates how the IPD framework enables researchers to overcome this limitation by generating out-of-sample estimates for periods lacking DXA data (such as 2021-2023) that closely approximate what we might find if DXA measurements were available. This approach allows for valid inference that remains aligned with fundamental research questions despite discontinuities in measurement availability.

Implementing this IPD correction requires a small subset of “gold standard” labels in addition to “predicted” labels for the outcome of interest. Within this framework, we designate DXA-measured whole-body fat percentage as the preferred measure because it directly quantifies body composition, while we consider BMI the “predicted” measure, as it serves merely as a proxy for adiposity. We leverage the relationship between DXA and BMI-measured adiposity across covariates of interest from the 2011-2017 period to estimate a bias term for each predictor. This bias term then adjusts the inference parameters derived from BMI-only labeled data in later NHANES waves (2021-2023) where our preferred DXA measure is unavailable. Essentially, this approach enables research continuity and consistent population inference across domains with heterogeneous measurement availability.

## Analysis

First, we begin with the 2011-2017 NHANES data, where BMI, WC, and DXA measures are collected for each respondent. We demonstrate how these three measures are correlated, but with varying levels of disagreement for different attributes.

Next, we specify three sets of univariate logistic regressions, each set with a different measure of obesity as the binary outcome of interest and the same categorical measures of self-reported sex, race, age, education, household income and smoking status as predictor variables.^27^ The only difference between the three sets of models is which measure for adiposity is used to classify the binary obesity outcome - BMI, WC or DXA. To interpret these coefficients as odds ratios, we exponentiate them.

It is important to note that for this in-sample portion of the analysis, we have complete data for all three obesity measures (BMI, WC, and DXA). To illustrate how progressively more missing DXA data biases our results, we artificially create missing data scenarios by randomly designating certain proportions of DXA measurements as “missing”. This controlled experiment allows us to quantify the impact of missing data on population level obesity estimates before applying our correction to genuinely missing DXA data in the 2021-2023 NHANES survey cycle.

Then we calculate IPD-corrected estimates using a randomly selected hold-out set of labeled data for the PPI++ procedure outlined in Angelopolous 2024 and implemented in the ipd R package available on CRAN.^24, 35^ We use this as a validation exercise to show that the IPD corrected estimates approximate the preferred DXA estimates, relative to the BMI estimates. We use a 70%-30% labeled-unlabeled split to demonstrate the efficacy of our IPD correction, as that is roughly the proportion of unlabeled to labeled data we have for the 2021-2023 out-of-sample analysis. In general, a larger proportion of labeled data will better approximate the “preferred” measure. We show results from a 50%-50% and 30%-70% split in the Appendix **??** to demonstrate how the correction may perform if applied to data contexts with different proportions of labeled and unlabeled data. This is one of the benefits of setting up this in-sample controlled experiment before moving to out-of-sample estimates.

Finally, we move to the 2021-2023 NHANES data for out-of-sample estimation with IPD correction, as these years do not contain DXA measures of body fat. Here, unlike our controlled experiment with artificial missing data, we face genuine absence of DXA measurements – a common data limitation in most large-scale obesity studies. We perform the PPI++ correction on the entire 2011-2023 data, with the 2011-2017 data serving as our “labeled” data ground truth for calibration and the 2021-2023 data being our “unlabeled” subset as described in Angelopolous 2024.^24^ The PPI++ correction enables us to efficiently calibrate BMI-based inferences and reflect the appropriate level of uncertainty that BMI introduces when it is used as a stand-in for DXA-based directly measured adiposity.

### NHANES 2011-2017 in-sample validation

In this section we highlight two primary conclusions from our in-sample analysis with BMI, WC and DXA-based measurements of adiposity. First, we definitively demonstrate that BMI is not “good enough” as a measure of adiposity for population-level obesity research. For example, consider how for males, the shape of the BMI and DXA distributions more similar than the distributions for females. Several people are classified as obese according to one measure, but not the other and vice versa. These differences for males and especially females are further exacerbated when looking at more particular social categories, such as those who identified as black or for those who responded having less than a high school education. Similar plots for all levels of each obesity measure and covariate level can be found in the supplementary materials. These prediction misclassifications lead to substantially biased estimates of obesity associations for different demographic groups. This finding challenges the widespread assumption that BMI provides adequate approximation for adiposity in population studies.^36^

Second, the inference with predicted data (IPD) statistical correction performs well in practice, shrinking this bias to generate valid estimates that closely approximate the effect size estimates we would obtain using the “gold standard” direct measure of adiposity. In our regression analyses using data from 2011-2017, we found that both BMI and WC-measured obesity yield conclusions about predictor associations that yield opposite results from those produced by the ground truth DXA measure of adiposity, underscoring the critical importance of our correction method for accurate obesity research.

Critically, the magnitude and direction of the associations between race and obesity depend on which measure of obesity is used. For example, consider the obesity odds ratio for the black racial/ethnic group relative to white. According to the WC measure, the estimate for black respondents is not statistically significant (OR: 1.0, 95% CI: [0.89,1.20]). Using BMI, we estimate a significantly higher odds of being labeled obese (OR: 1.43, 95% CI: [1.22,1.68]) and for DXA, sign flips, black respondents have lower odds of being labeled obese, though it is not statistically significant (OR: 0.90, 95% CI: [0.77,1.05]). The in-sample validation exercise using the 2011-2017 NHANES data allows us to compare the IPD corrected estimates with the BMI, WC and DXA estimates. For a given proportion of labeled and unlabeled data (blend of BMI and DXA data, in this case), we evaluate whether this conclusion about race aligns more with the BMI conclusion, WC conclusion or the DXA conclusion. With reference to the 70%/30% labeled-unlabeled split used to produce the IPD estimate in Figure 3, we see that IPD-adjusted BMI estimates closely approximate the DXA-based estimates. For example, for black respondents, the IPD-adjusted BMI odds ratio estimate is 0.91 compared to the DXA-based estimate of 0.90. This validation exercise demonstrates how BMI obscures an important association that would be observed when using a more direct measure of adiposity.^8, 37, 38^ The magnitude and direction of the bias, as well as the amount of uncertainty introduced by using BMI rather than a more precise measure of obesity, will depend upon both the population and covariates of interest.

**Figure 3:**
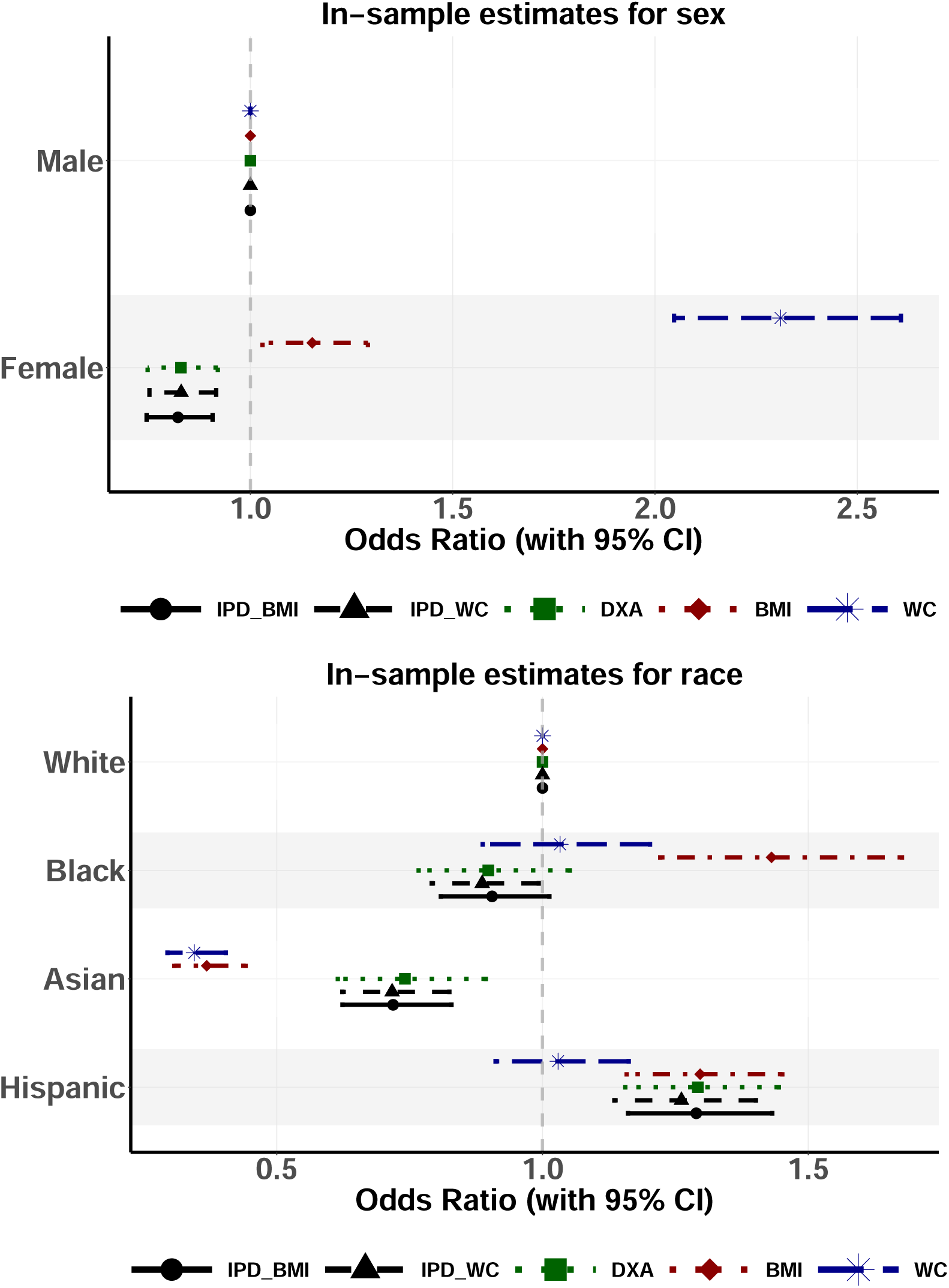
Odds ratios estimated using BMI, WC and DXA-measured obesity vary in effect size, direction and statistical significance for the same observed covariates. Compared to BMI and WC, the IPD corrected estimates more closely resemble DXA, which directly measures adiposity. These estimates are performed using 10-year adjusted survey weights and a 70%-30% labeled-unlabeled split (see the Appendix A.1, A.7 for more detail). Similar plots for all covariates can be found in A.3.

### NHANES 2021-2023 out-of-sample estimation

Now, when we evaluate the out-of-sample associations using IPD corrected estimates instead of the BMI or WC estimates we approximate the association we might estimate if the COVID-19 pandemic had not interrupted the collection of DXA measurements in 2021-2023. Without observing DXA we cannot know for sure, but the in-sample validation from Figure 3 gives us confidence in the 2021-2023 results. The 2021-2023 IPD corrections we estimate are shown in Figure 4. Notably, the BMI and IPD corrected estimates disagree about the sign and significance for black respondents (BMI OR: 1.47, 95% CI: [1.14,1.90]), (IPD corrected OR: 0.90, 95% CI: [0.82,1.0]) and female respondents (BMI OR: 1.09, 95% CI: [0.95,1.24]), (IPD corrected OR: 0.82, 95% CI: [0.76,0.89]). Even in the absence of DXA data, we should conclude that female respondents are at a significantly decreased odds of being labeled as obese in the 2021-2023 NHANES sample since we used the whole data, even though we would conclude the opposite if solely considering BMI, or WC for that matter.

**Figure 4:**
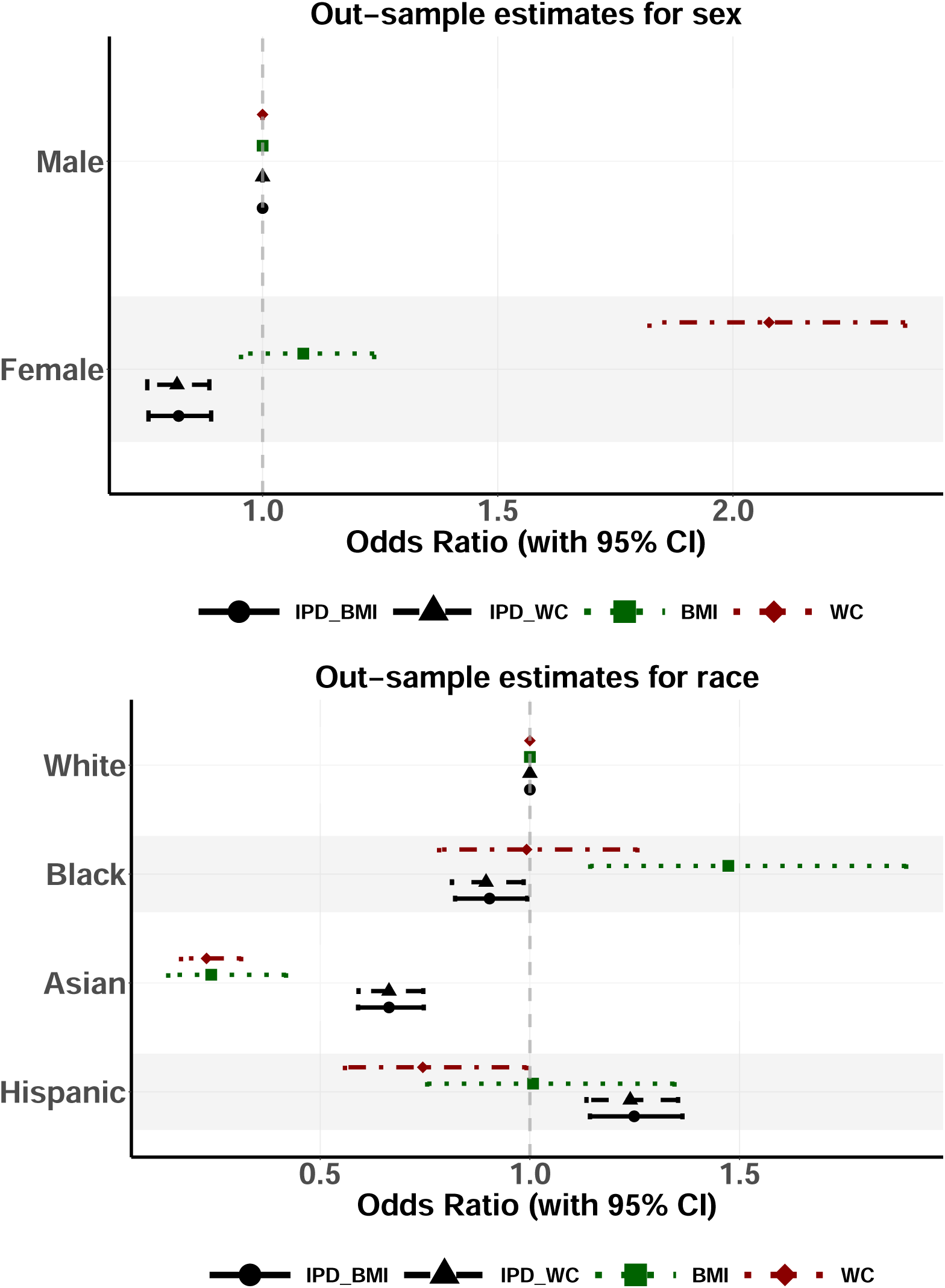
Whole-body fat percentage measured using a DXA scan is still the conceptually preferred measure adiposity for obesity research. Only anthropometric measures BMI or WC were collected by NHANES in 2021-2023 data due to interruptions from the COVID-19 pandemic. The IPD corrected estimates for BMI and WC approximate the association between DXA adiposity and observed covariates had DXA been available. This is estimated using observed outcomes for BMI, WC and DXA in the 2011-2017 sample where all three measures were collected for the same individuals.

Anthropometric measures like BMI or waist-to-hip ratio measure something, but they do not measure adiposity. Yet, cutting-edge obesity research continues to rely on these proxy measures.^39^ Our analysis demonstrates that this reliance on BMI for population-level inference leads to significantly biased estimates for associations between obesity and common predictors, particularly for demographic subgroups such as black respondents, female respondents, and respondents with less than High School education.

The problem is not that BMI should not be used at all – we recognize the practical reality that DXA scans and other direct adiposity measurements are often unavailable due to cost constraints or data collection limitations, as exemplified by the absence of DXA data in NHANES 2021-2023. Rather, the critical issue is that when researchers use BMI as a proxy for adiposity without appropriate statistical correction, they risk drawing incorrect scientific conclusions that may ultimately misinform public health policy.

Our findings reveal substantial divergence between BMI-based and DXA-based estimates of obesity associations across key demographic and socioeconomic factors. This divergence is not merely academic – it has real implications for how we understand obesity epidemiology and potentially for how resources are allocated in addressing health disparities.^40, 41^

The IPD framework offers epidemiologists and public health researchers a practical solution to this measurement challenge. By leveraging the relationship between BMI and DXA measurements in available data (such as NHANES 2011-2017), researchers can generate corrected estimates that closely approximate what would be found if the “gold standard” measurement were available across all time periods. Our validation results demonstrate that this correction performs well in practice (Figure 3). Indeed, IPD is a useful tool in general if you are missing data and assume minimal change over time.

We encourage investigators to be precise in their terminology—using “BMI” or “body proportion” in scientific writing when those are the actual measurements employed, rather than terms like “fat,” “body composition,” or “adiposity” that imply more direct measurements (see Perry and Ciciurkaite’s work for a nice example).^42^ More importantly, we urge researchers to consider statistical correction methods like IPD when working with proxy measures that are conceptually misaligned with the true variable of interest which requires including a small subset of gold standard data to calibrate on.

This approach extends beyond obesity research to other domains where screening tools (e.g., blood pressure readings or depression questionnaires) are often mistakenly equated with the conditions they screen for. By implementing appropriate statistical corrections, researchers can perform valid inference even when limited to imperfect proxy measures, thereby improving the validity of estimates and more responsibly informing public health interventions.

## Limitations

First, we must reiterate that no empirical measure of obesity is “true,” as the very definition of obesity – “excessive adiposity that presents a risk to health” – lacks biological precision.^43^ This definition raises fundamental questions: Excess relative to what baseline amount of adiposity? How much of an increased risk is concerning? And crucially, an increased risk to which specific health outcomes? These inherent ambiguities make any single measurement approach necessarily imperfect. Some measures are simply more conceptually aligned with the outcome of scientific interest. It is up to the domain expert to define this for themselves within their expertise.

Second, performing IPD corrections does require at least *some* “gold standard” data – by which we mean the measure which the researcher conceptualizes as a true measure of their variable of interest. The proportion of these “gold standard” measures that is necessary to recover valid estimates will vary depending upon the context, but in practice we recommend no less than 10%.

Finally, the analyses we present above rely on data collected in different years but with the same survey methodology. As it stands now, we would not be able to use labeled and unlabeled data collected in one domain to correct estimates for data collected in another using an IPD correction. This transportability issue is an important practical challenge that requires further attention from survey statisticians.

## Data Availability

All data produced are available online at https://wwwn.cdc.gov/nchs/nhanes/default.aspx

## Acknowledgments

AV gratefully acknowledges the resources provided by the International Max Planck Research School for Population, Health and Data Science (IMPRS-PHDS). We thank Amy Wang for excellent research assistance. We also thank Sidnee Moyer, Nicole Ward, Summer Shepherd, Kristen Olsen, Sarah Quinn, Patricia Louie for their feedback and suggestions.

## Author contributions

Adam Visokay - Literature search, conceptualization, formal analysis, accessed and verified the underlying data, project administration, validation, visualization, writing (original draft).

Kentaro Hoffman - Conceptualization, methodology, supervision, writing (review and editing).

Stephen Salerno - Methodology, software, accessed and verified the underlying data, writing (review and editing).

Tyler H. McCormick - Methodology, supervision, funding acquisition, writing (review and editing). Sasha Johfre - Literature search, supervision, funding acquisition, writing (review and editing).

## Generative AI disclosure statement

We utilized multiple Generative AI tools (OpenAI’s GPT-4 [including through GitHub Copilot]; Microsoft’s Copilot [based on the GPT-4 architecture]; and Anthropic’s Claude 3.5/3.7 Sonnet) in the production of this manuscript, in the following ways:

- Producing computer code for data cleaning and analysis
- Locating relevant research articles in the literature
- Brainstorming ideas and outlining the structure of the paper
- Proposing sentences to include in the manuscript
- Iteratively improving the concision and clarity of the writing

We have carefully reviewed all aspects of the manuscript for accuracy and coherence. All scientific insights, analysis and interpretation of data and scientific conclusions are made solely by the authors. All errors are our own. This disclosure is adapted from Professor Tyler Ransom.^44^

## Abbreviations

BMI: Body Mass Index
DXA: Dual-energy X-ray absorptiometry
WC: Waist Circumference
NHANES: National Health and Nutrition Examination Survey
OR: Odds-Ratio
IPD: Inference with Predicted Data
CRAN: Comprehensive R Archive Network
PPI++: Efficient Prediction-Powered Inference

## A Appendix

## A.1 Survey Weights

We follow the methodology outlined by the CDC.^45–49^ Despite the 2011-2023 spanning 12 years, the 2019-2020 survey cycle was canceled due to the COVID-19 pandemic. Therefore, we construct 10-year adjusted survey weights as there are only five 2-year cycles collected during the study period. We refrain from using data before 2011 because any DXA scan measures are imputed rather than measured directly. Due to how the covariates are operationalized from the raw survey responses we are missing a minority of observations for each variable (except for cigarette smoking where we focus only on those who responded *{*never, past, current*}*).

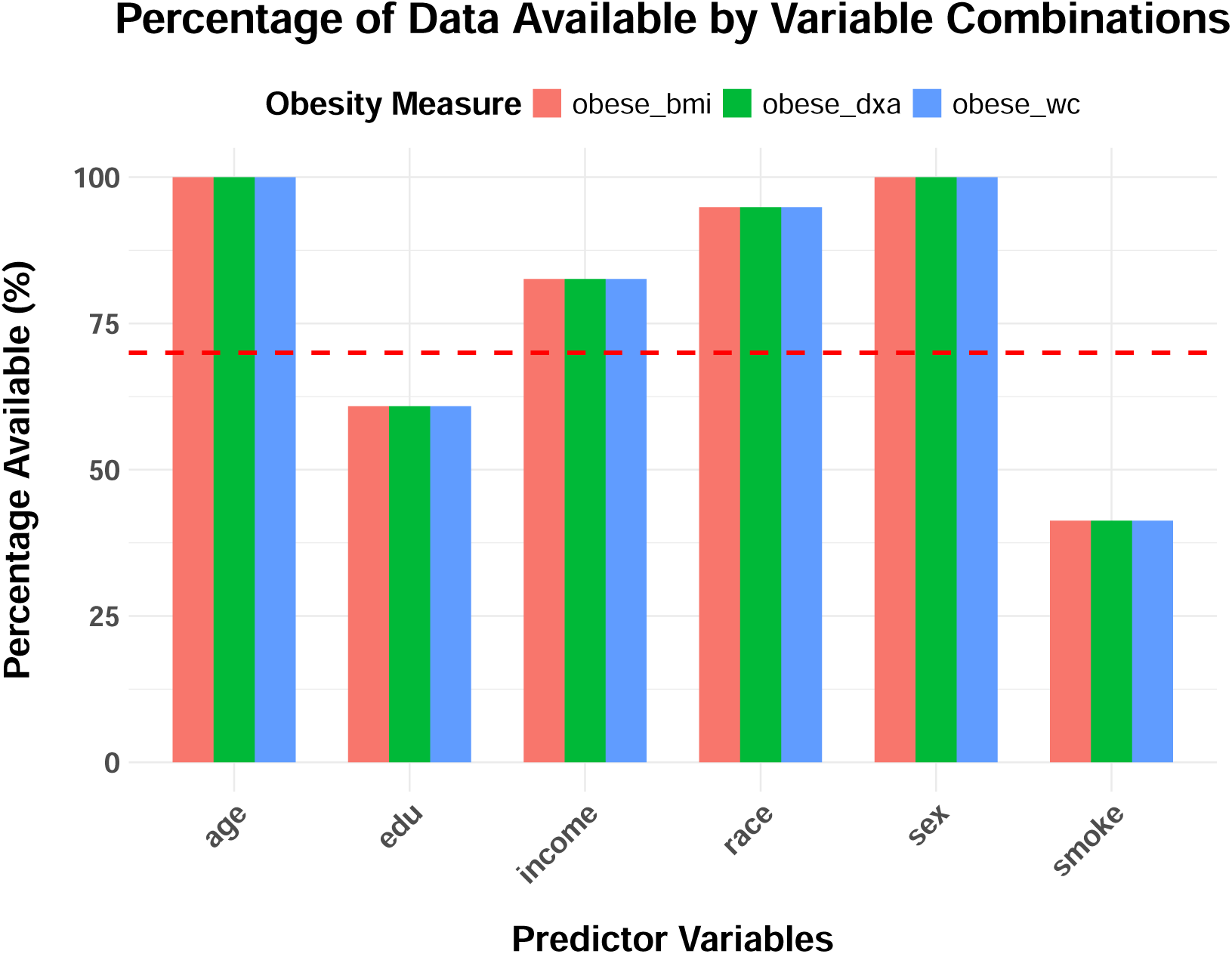

## A.2 Comparing distributions

Below are all of the overlapping distribution plots for each permutation of obesity measure (BMI, WC, DXA) and covariate (race, age, edu, income, smoker) by sex.

**Figure 5:**
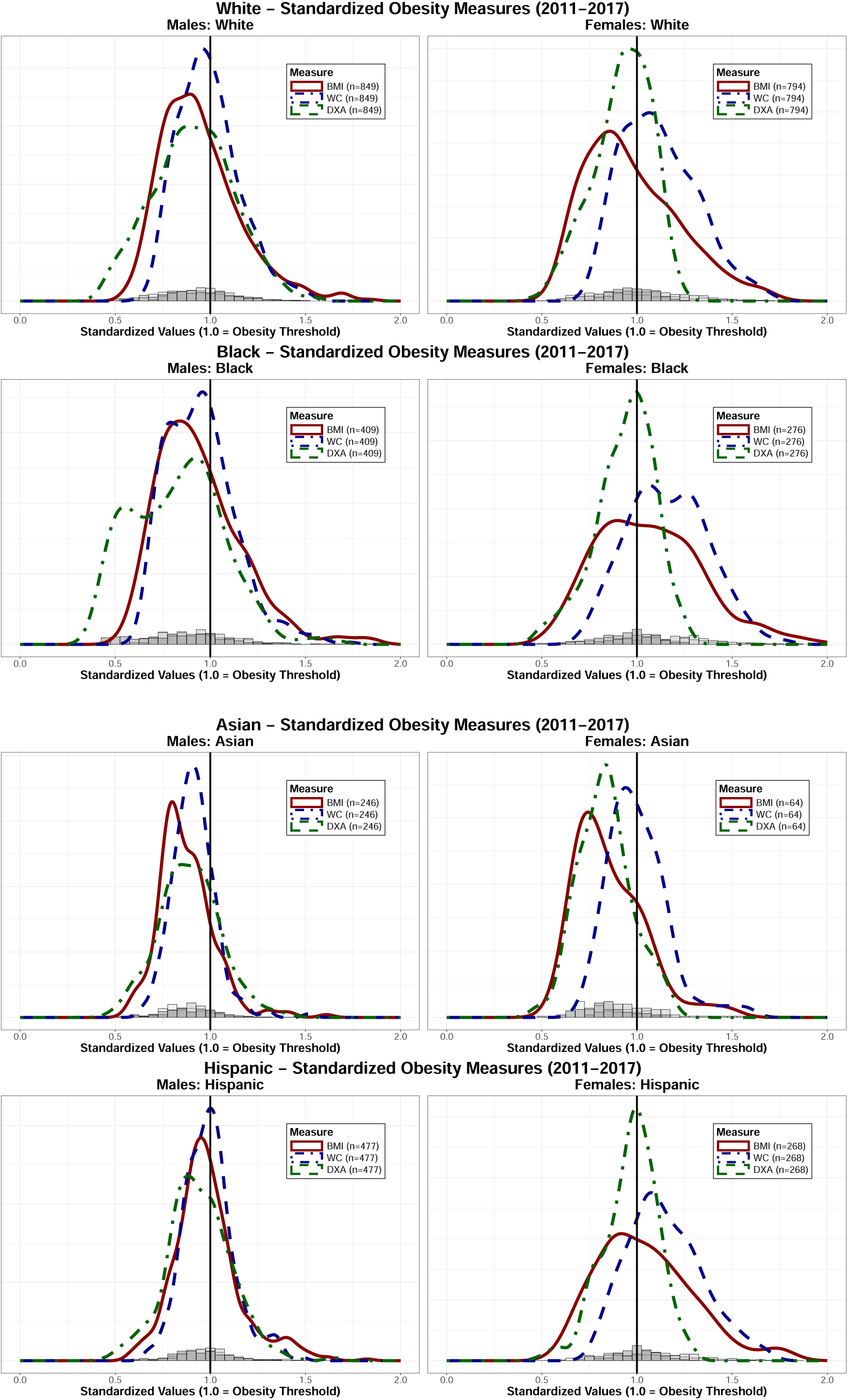
Race

**Figure 6:**
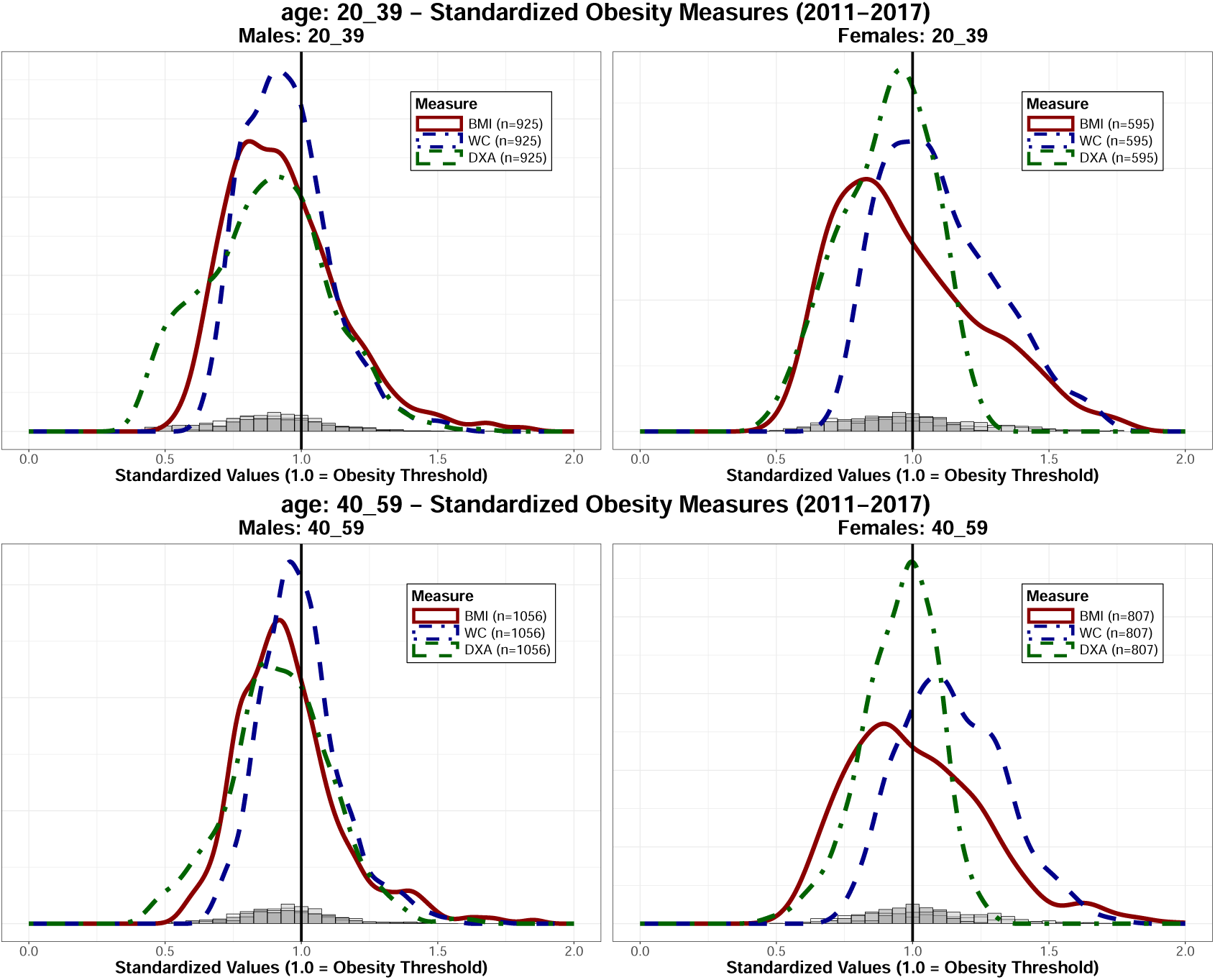
Age

**Figure 7:**
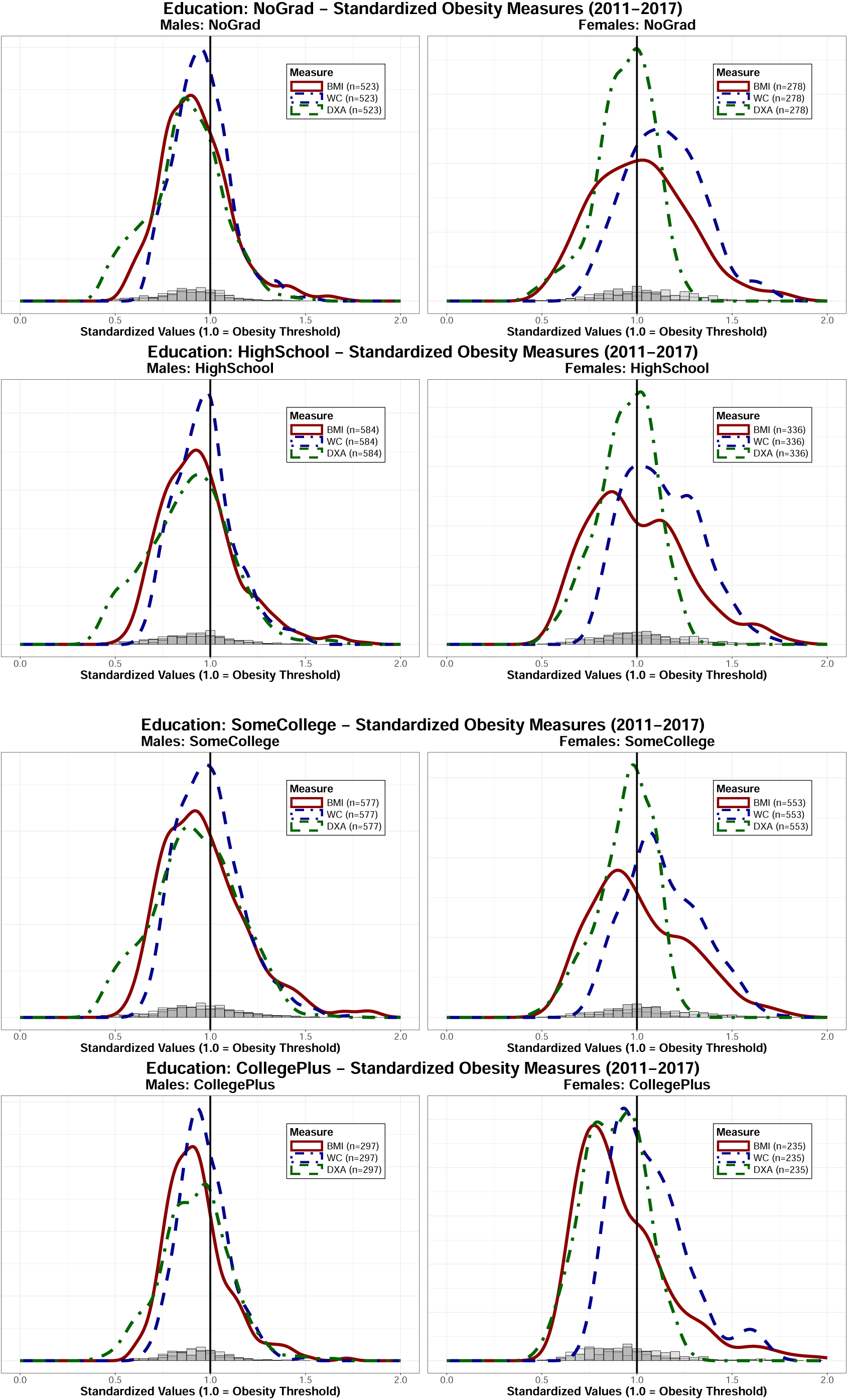
Education

**Figure 8:**
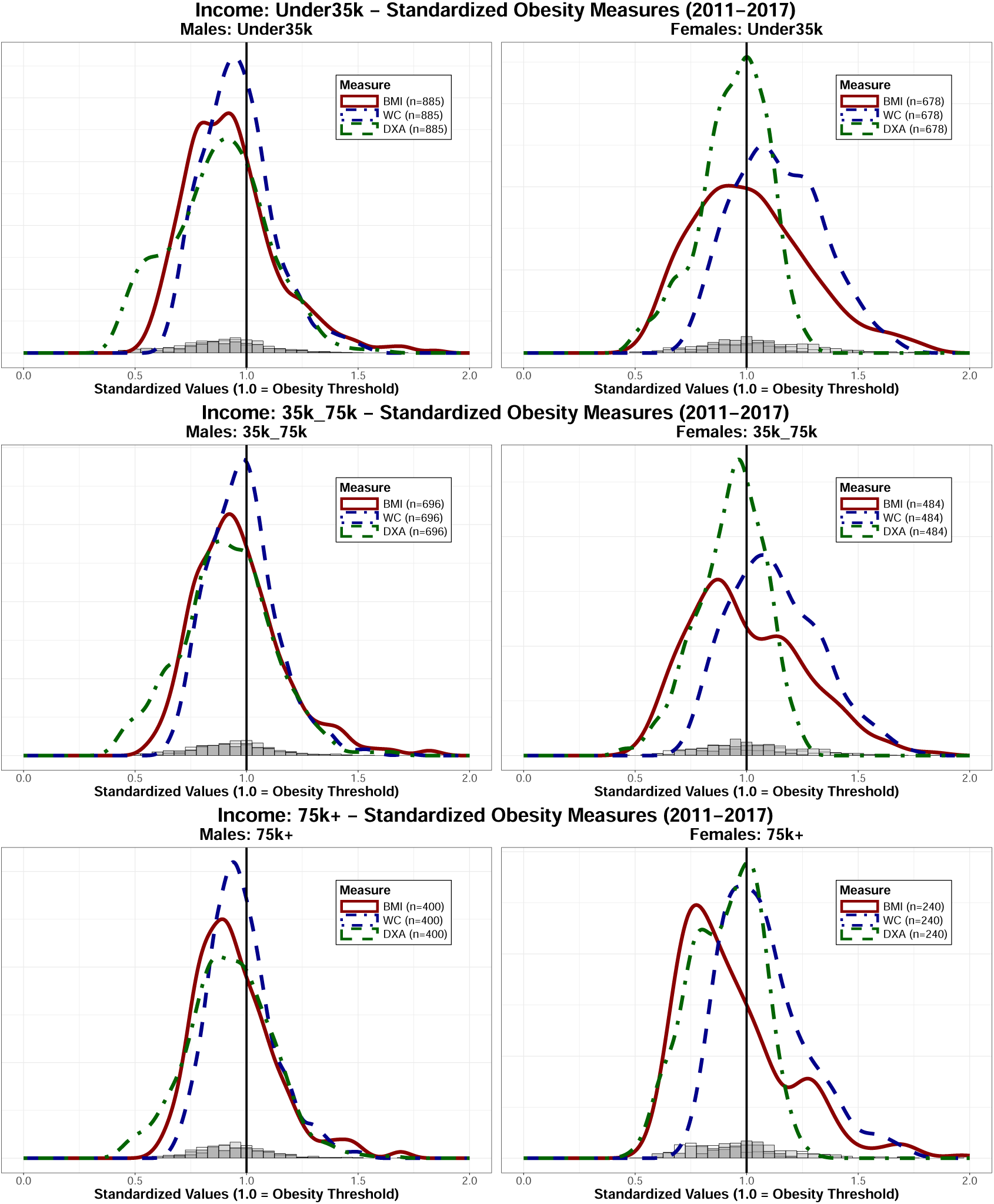
Income

**Figure 9:**
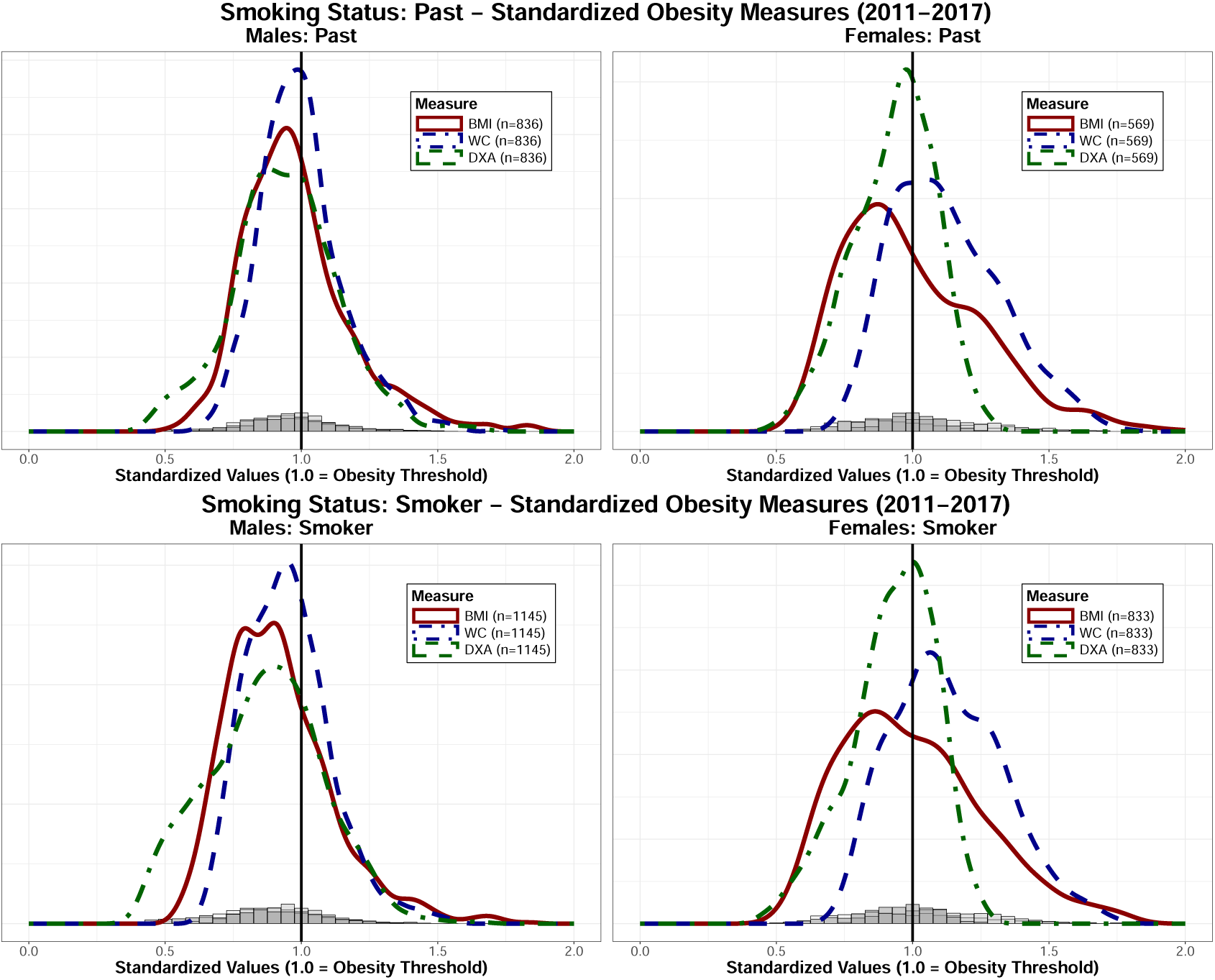
Smoker

## A.3 Full In-sample Results

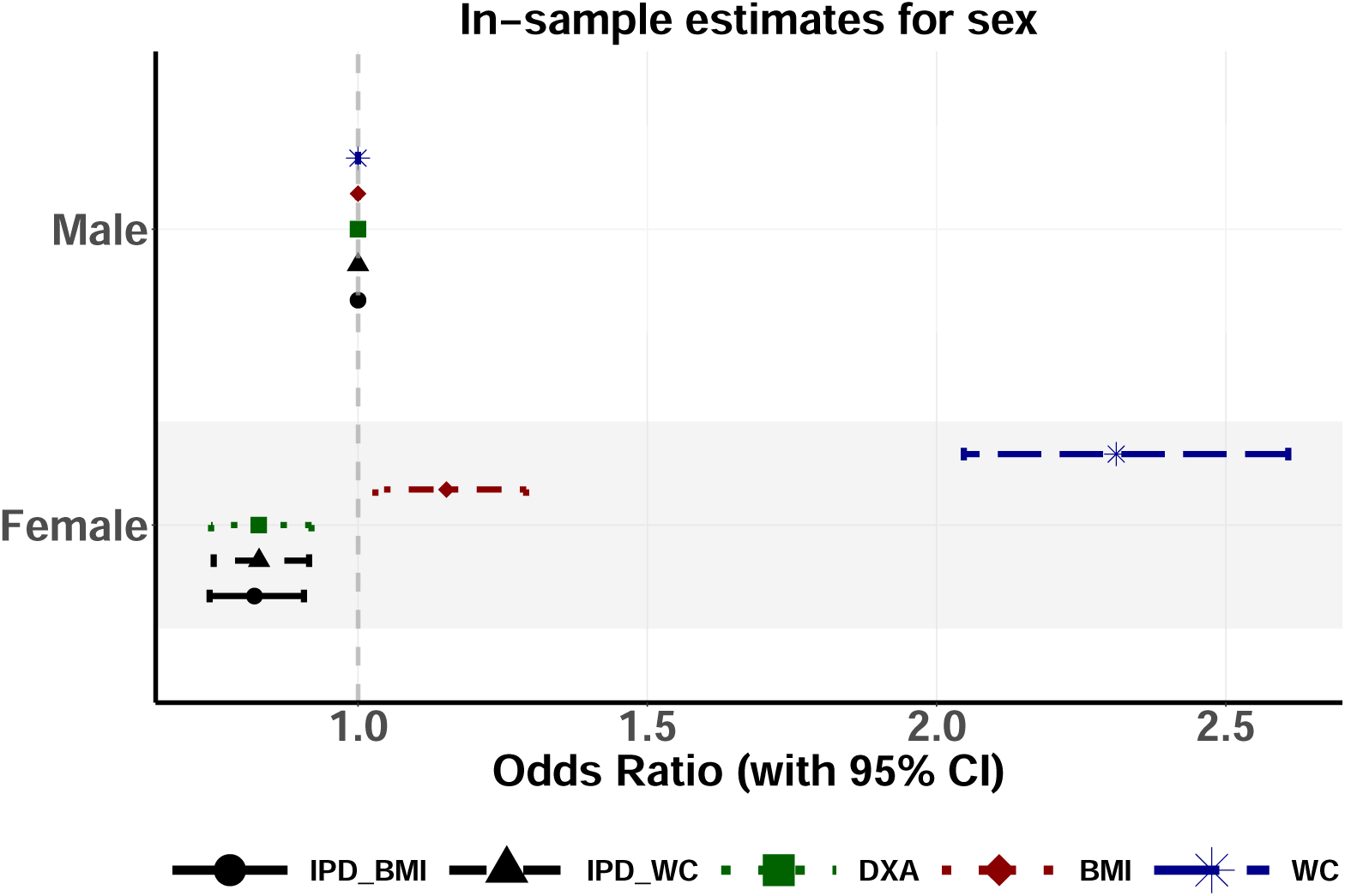
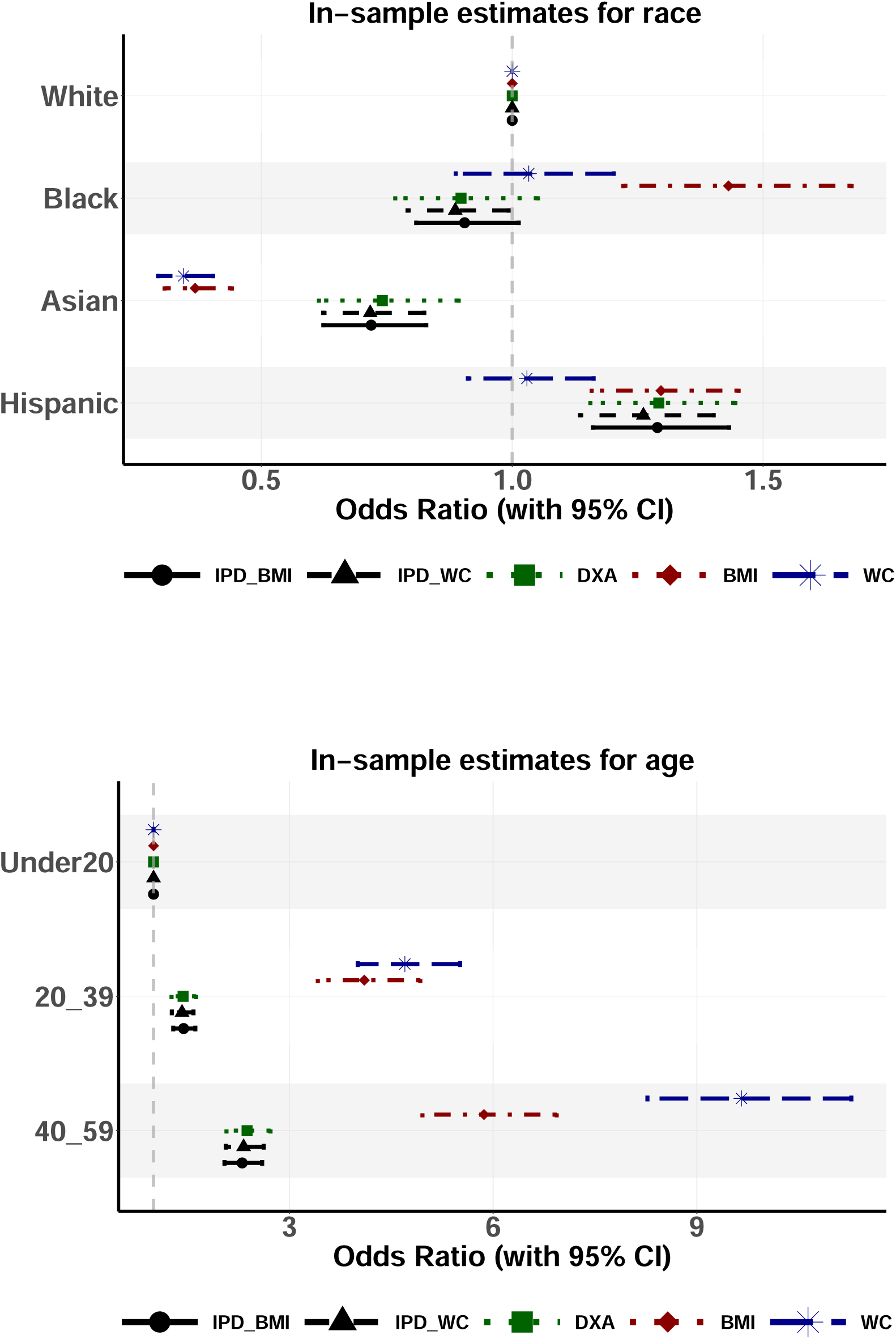
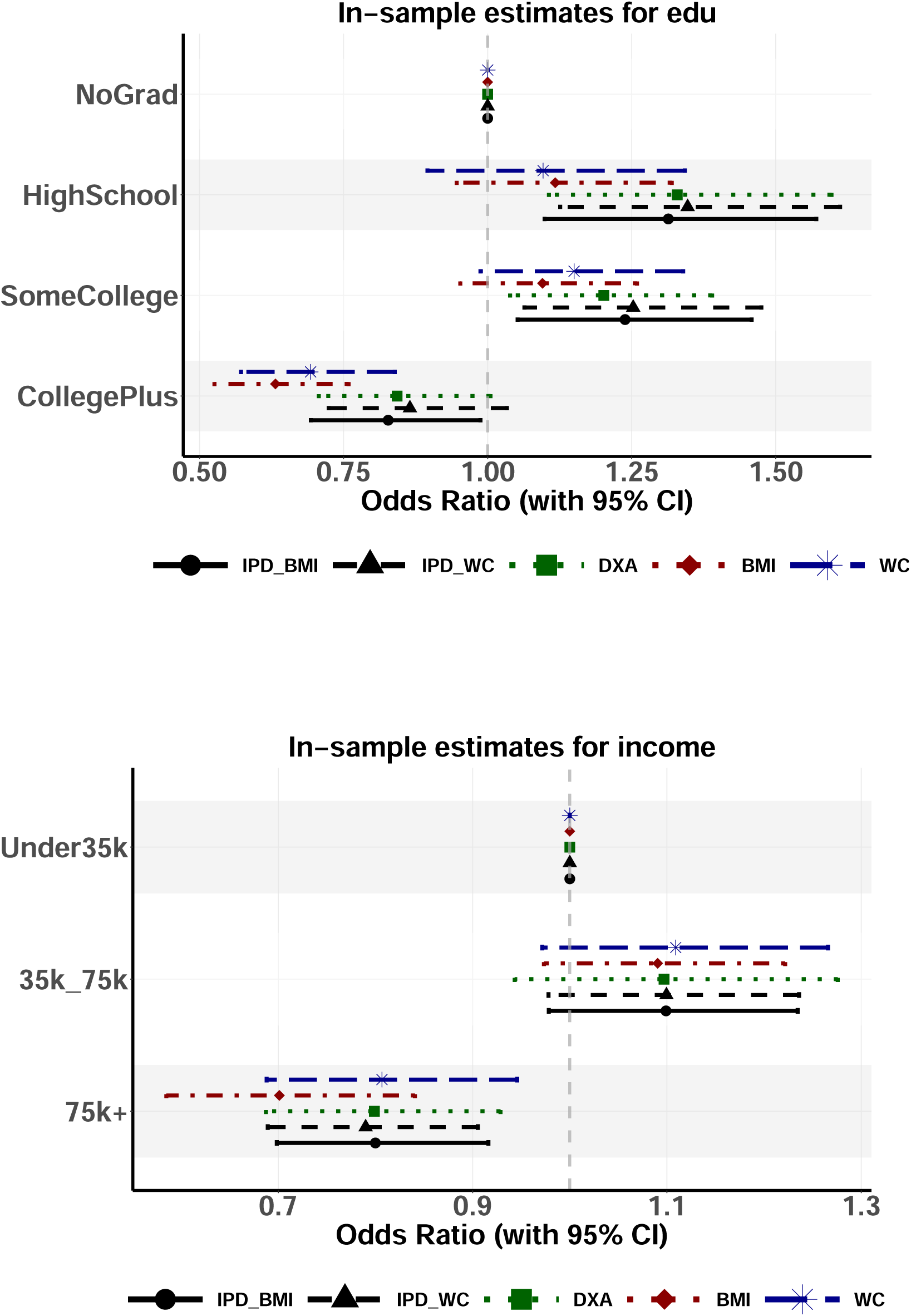
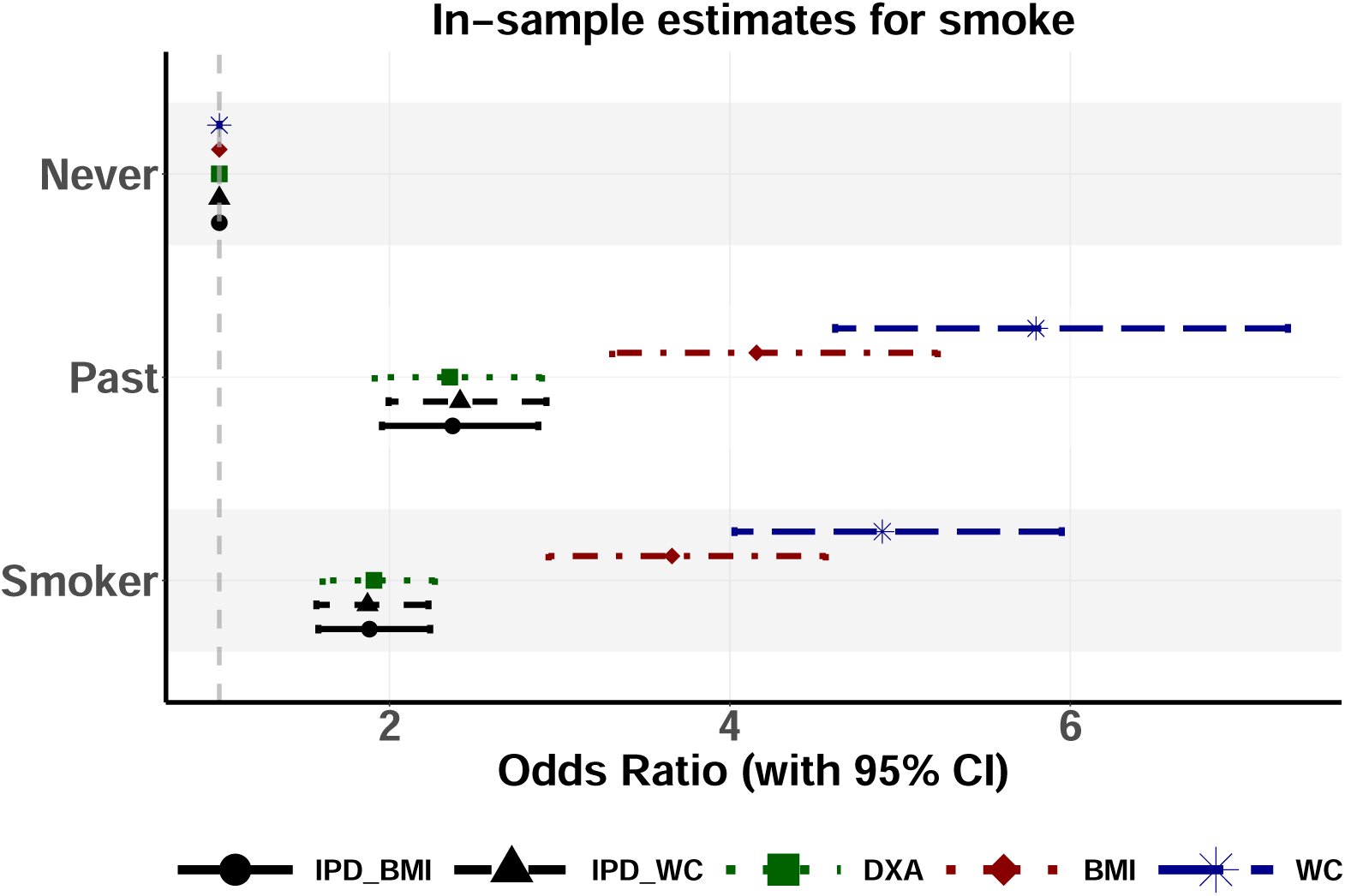

## A.4 Full Out-sample Results

We only include outsample results for sex, race and education due to missing data arising from changes to the NHANES collection procedure during the COVID-19 pandemic.

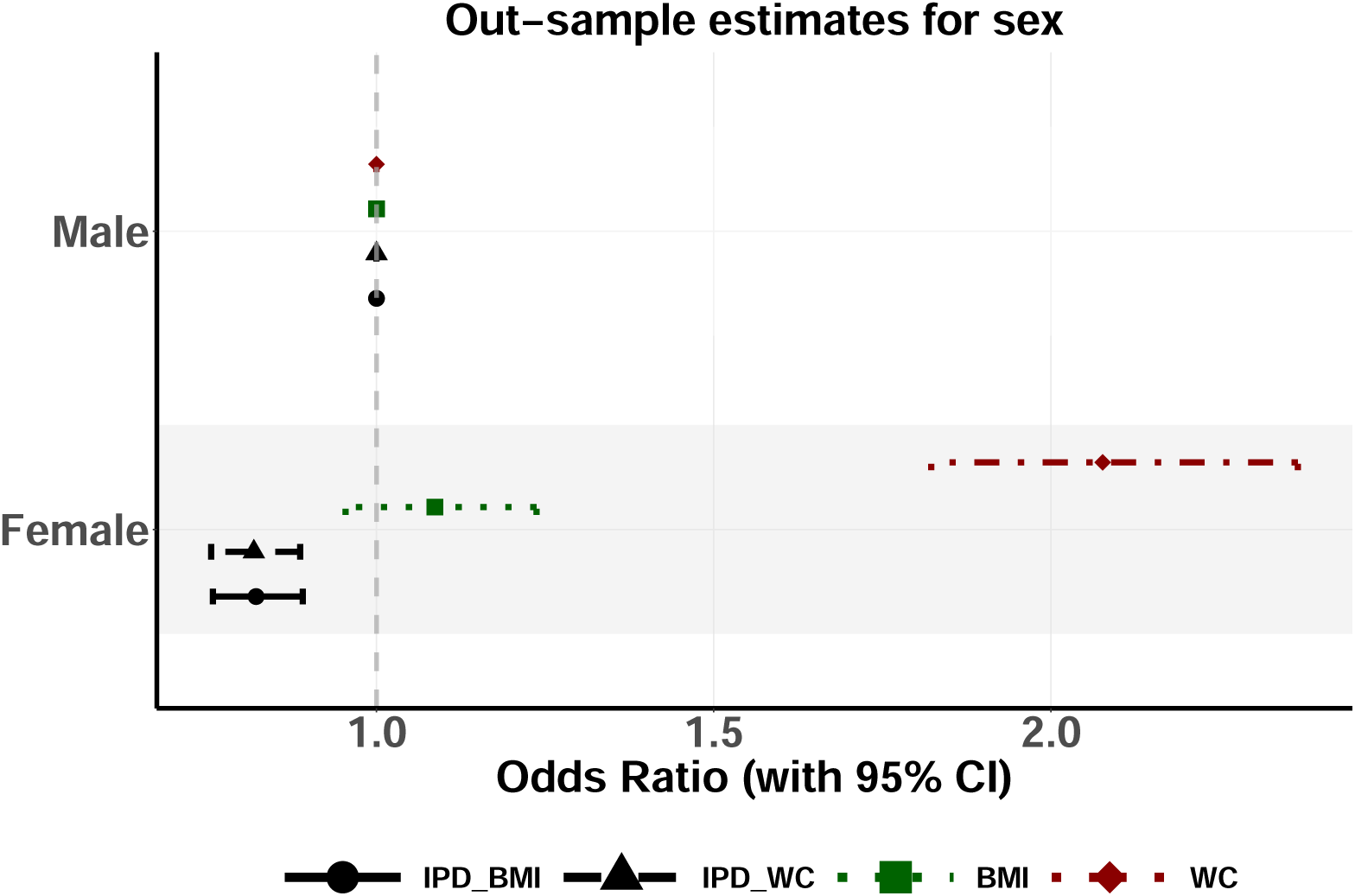
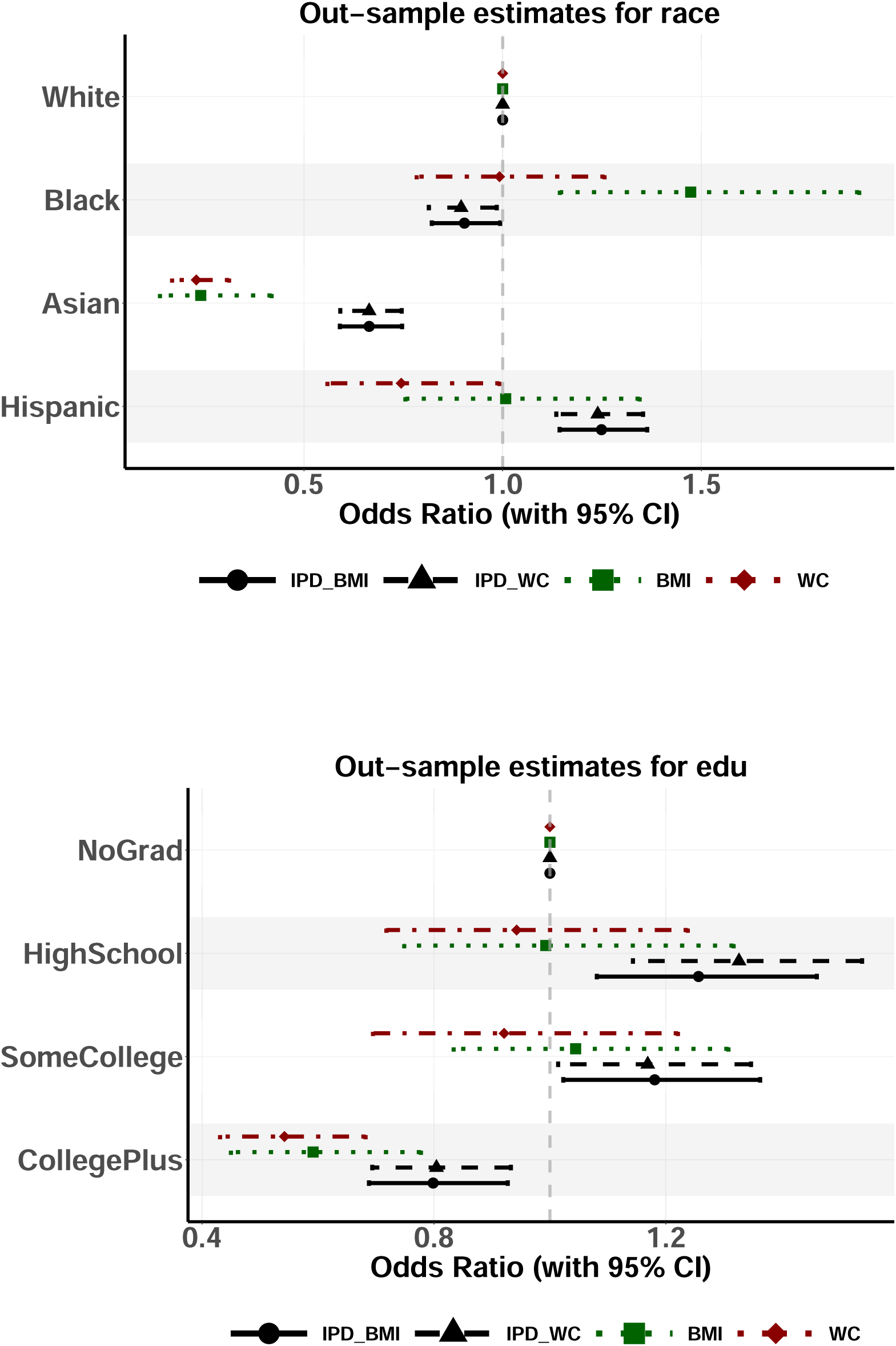

## A.5 Robustness Check: Continuous Obesity Outcomes

Below are estimates from a linear regression with continuous, standardized outcomes for BMI-score(kg/m^2^), Waist Circumference (cm) and DXA scan (total body fat percentage). This demonstrates that it is not simply a question of where one sets the obesity threshold(s) for categories - there is underlying variation in the population by demographic groups that arises from the continuous measures themselves.

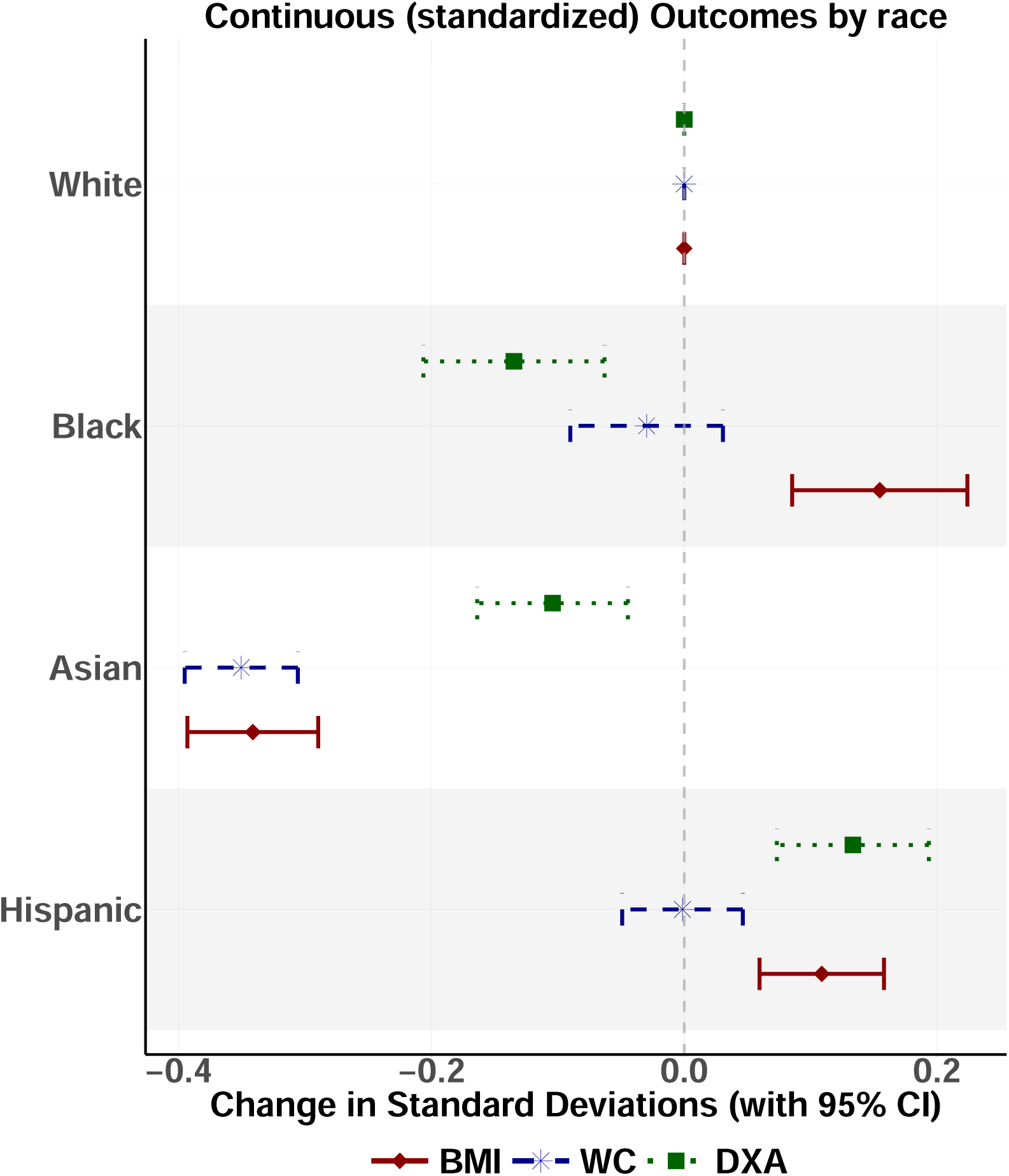
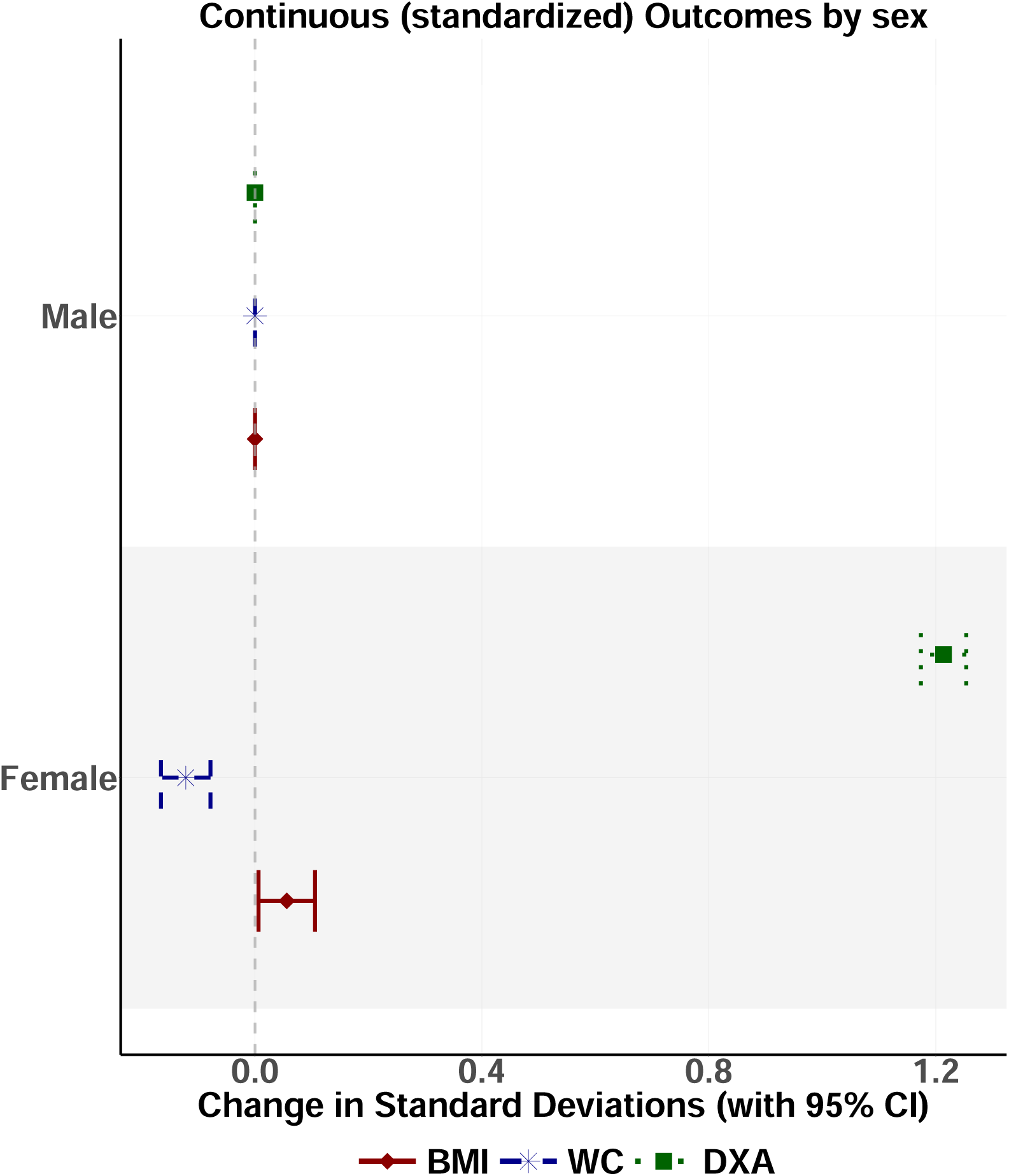
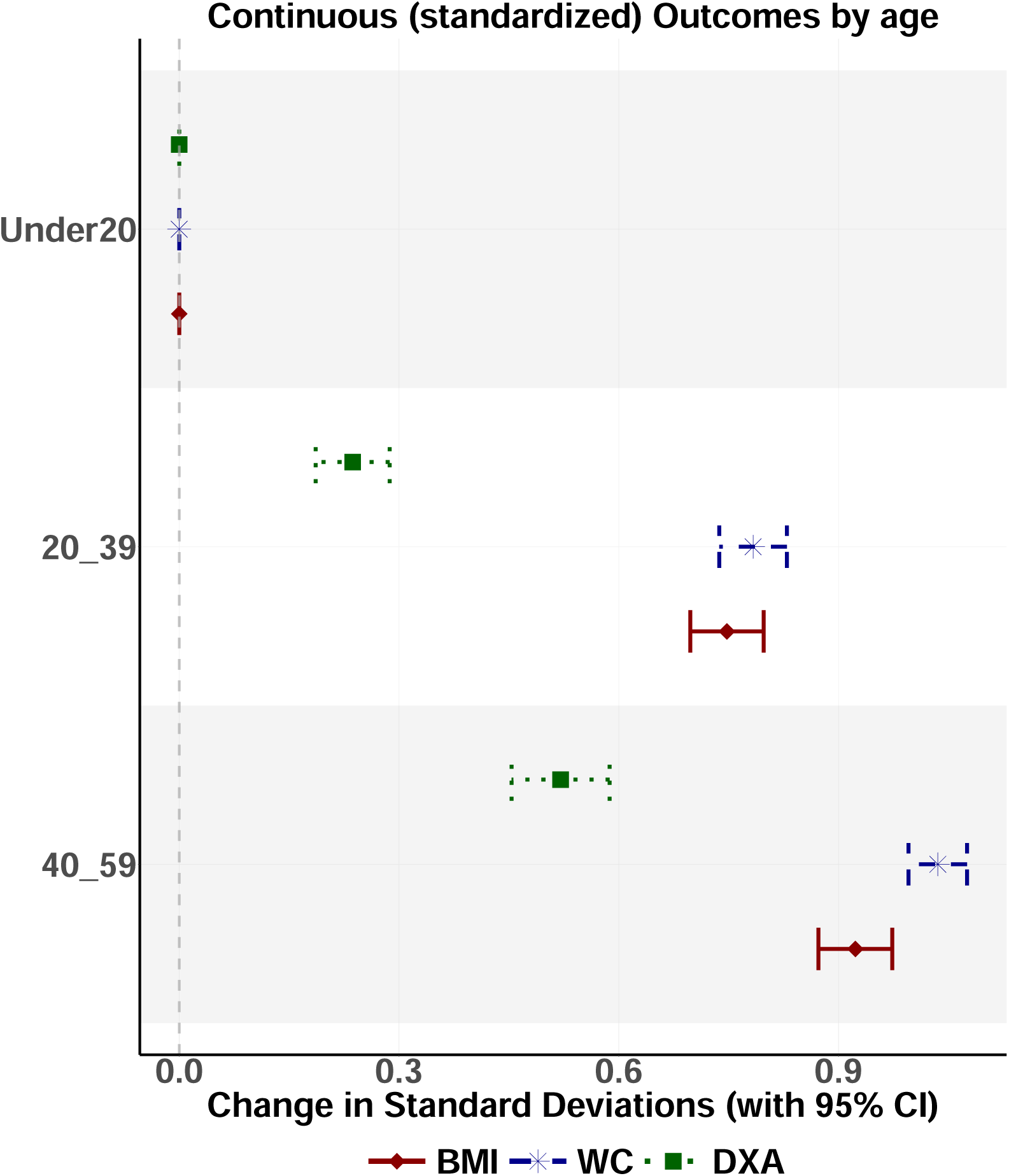
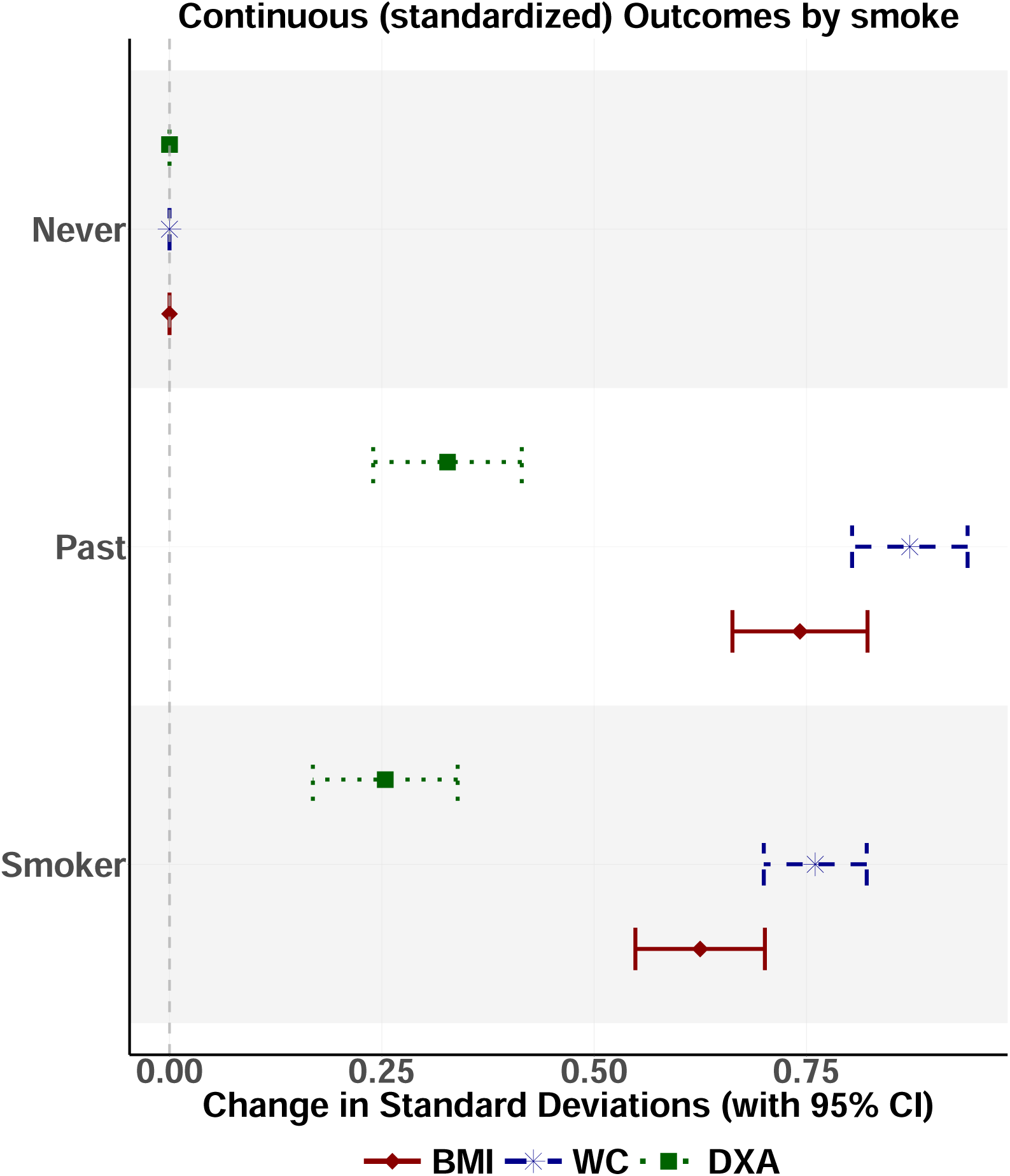
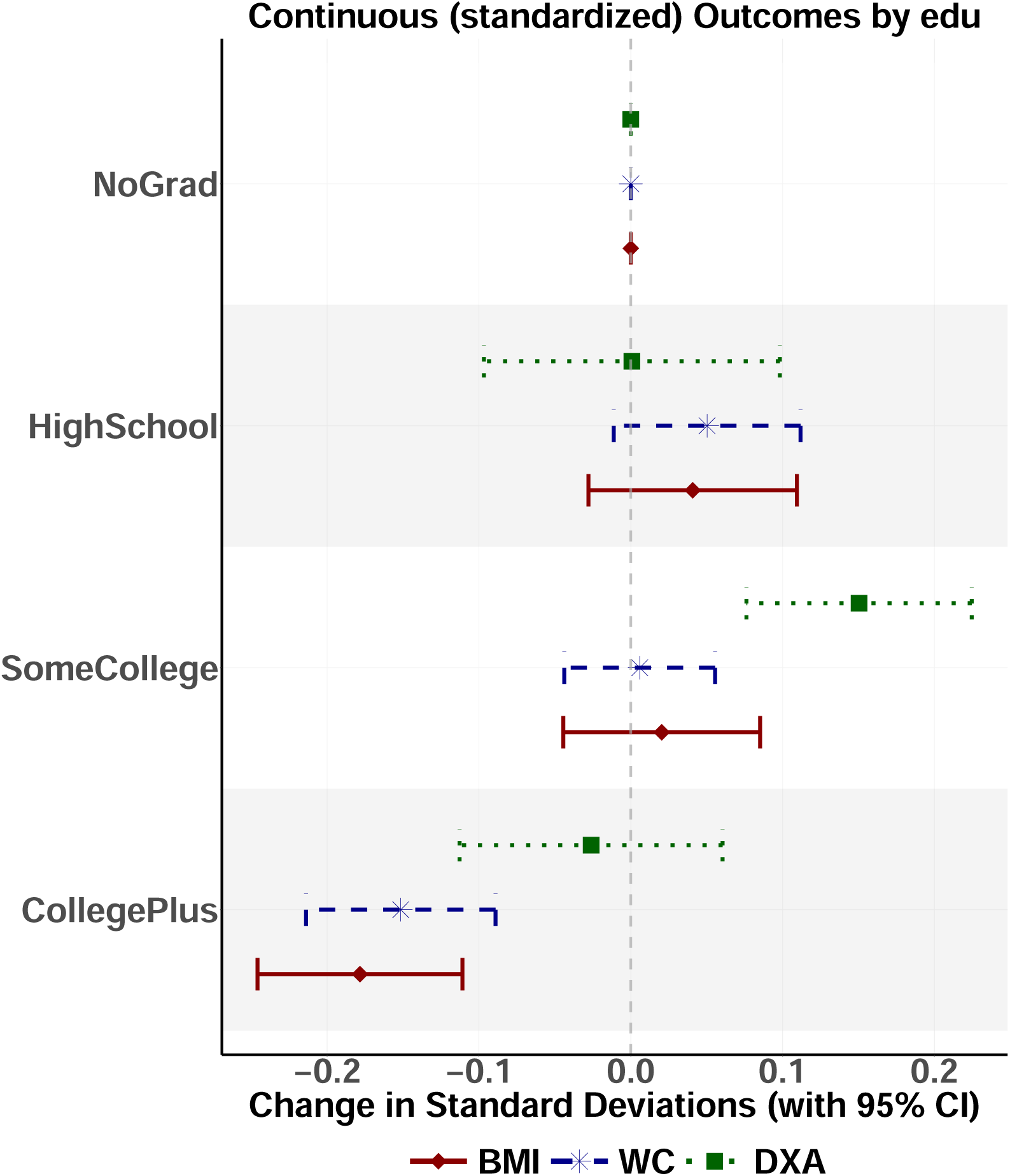
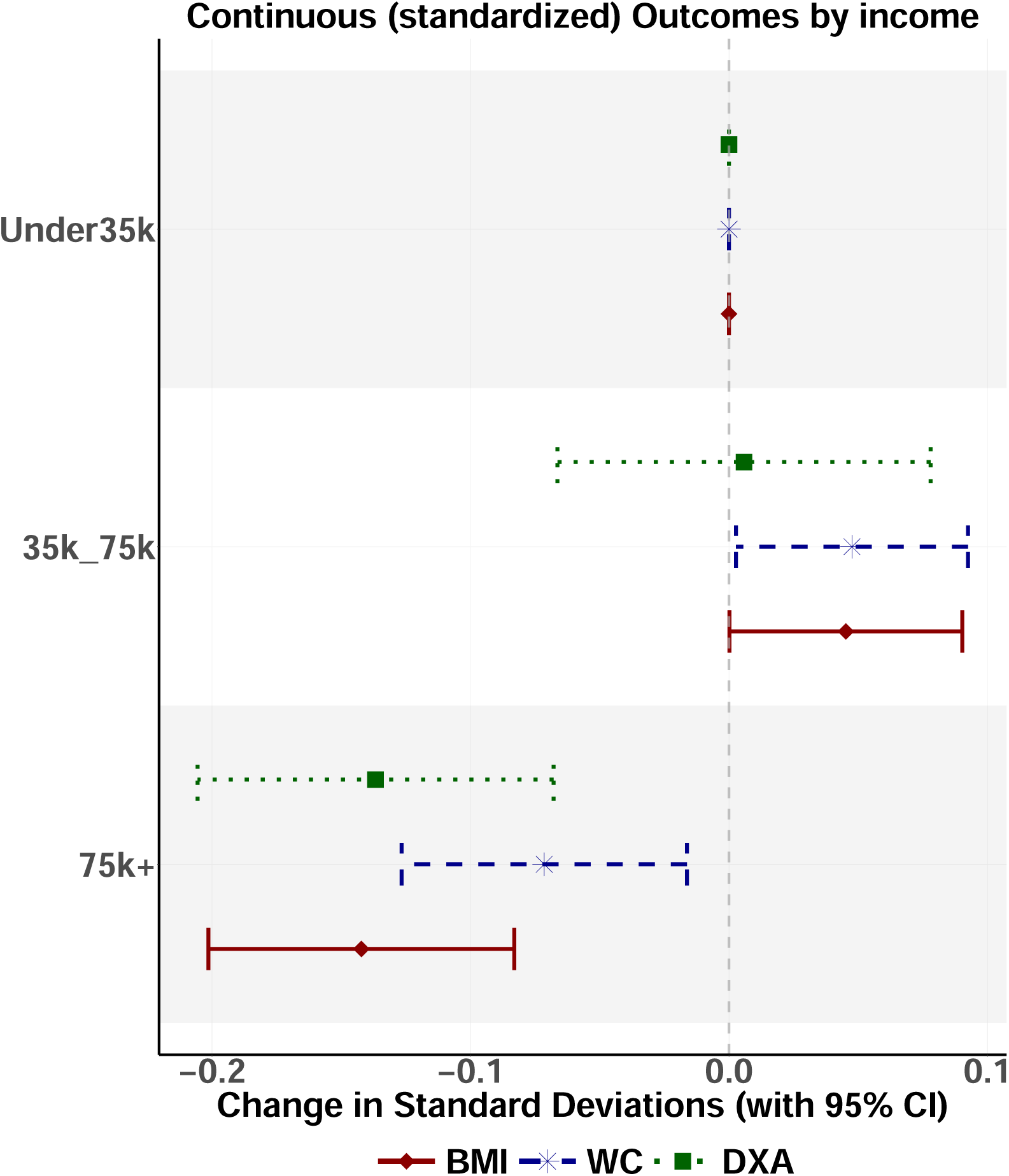

## A.6 Robustness Check: Alternative BMI Obesity Thresholds

Below are estimates from a logistic regression for Obesity as a binary outcome using different thresholds for BMI-score(kg/m^2^). Standard uses a BMI score of 30. BMI 1998 uses the original BMI categories from 1998 (27.8 for men and 27.3 for women). Ethnicity-specific cutoffs according to Caleyachetty et al. (28.1 for black, 30 for white, 23.9 for Asian and 30 for Hispanic) are explicitly designed to associate BMI with type 2 diabetes by race.^7^ The authors note that one limitation of these thresholds is they rely upon a 90.7% sample, but even

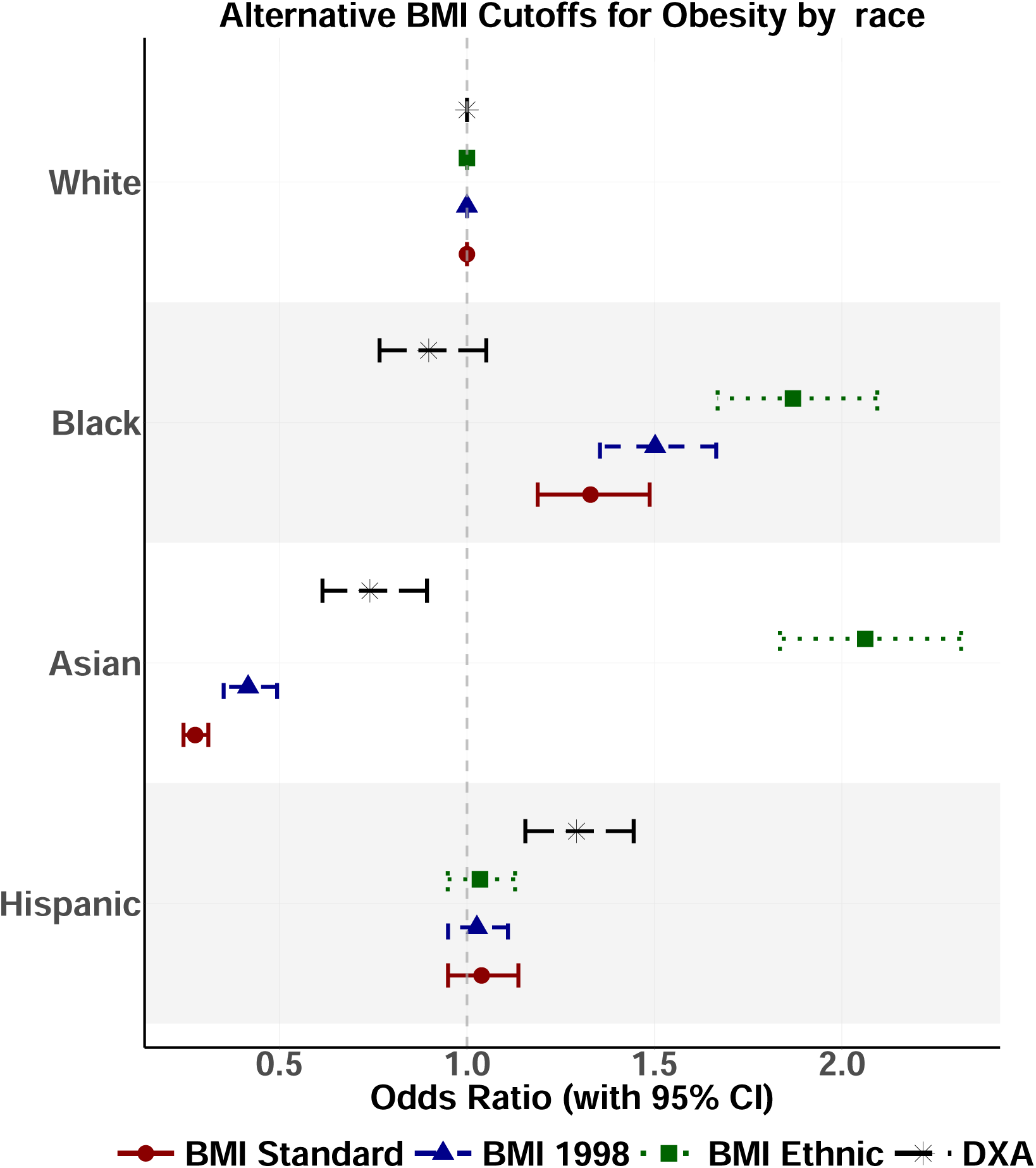
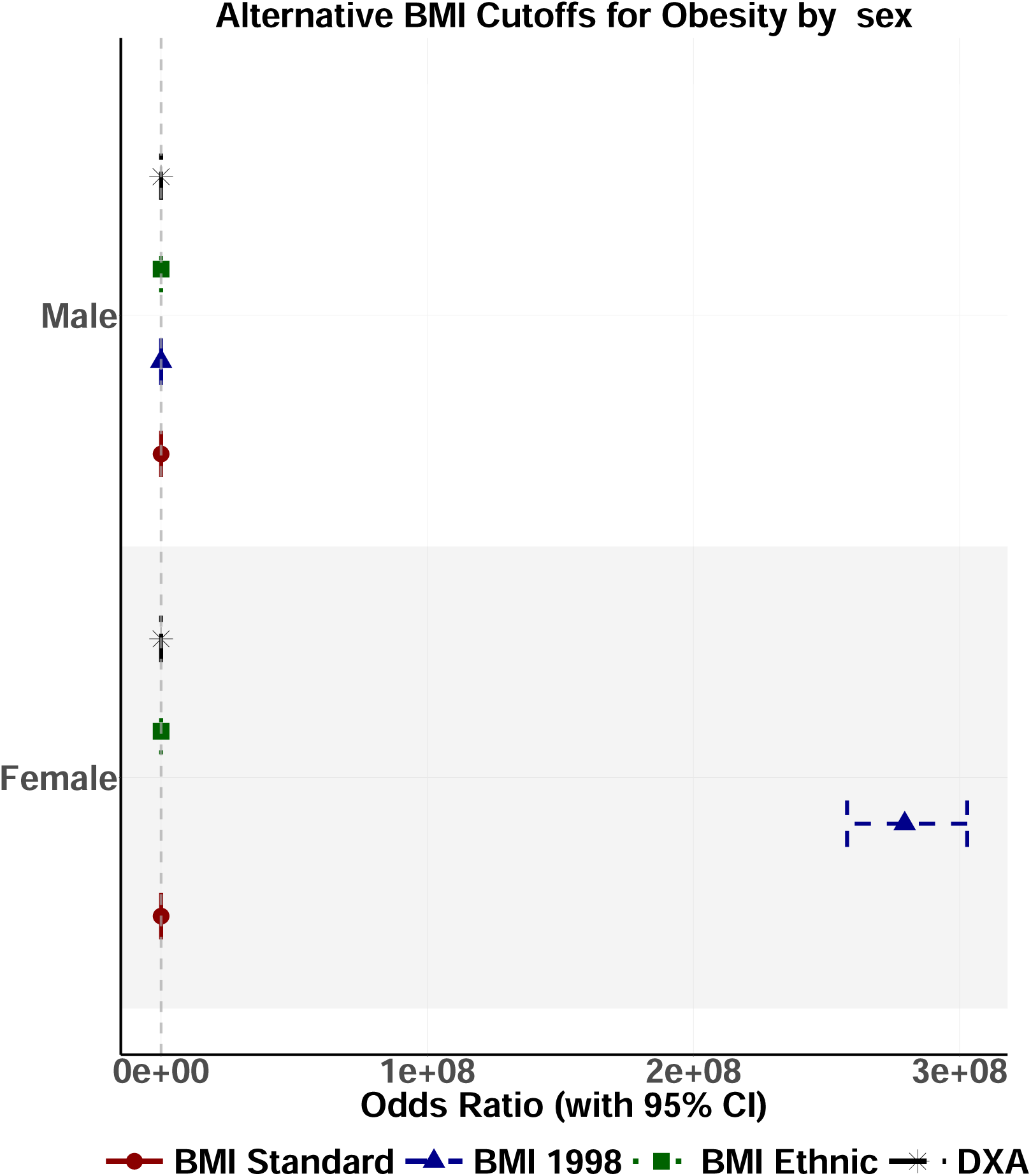
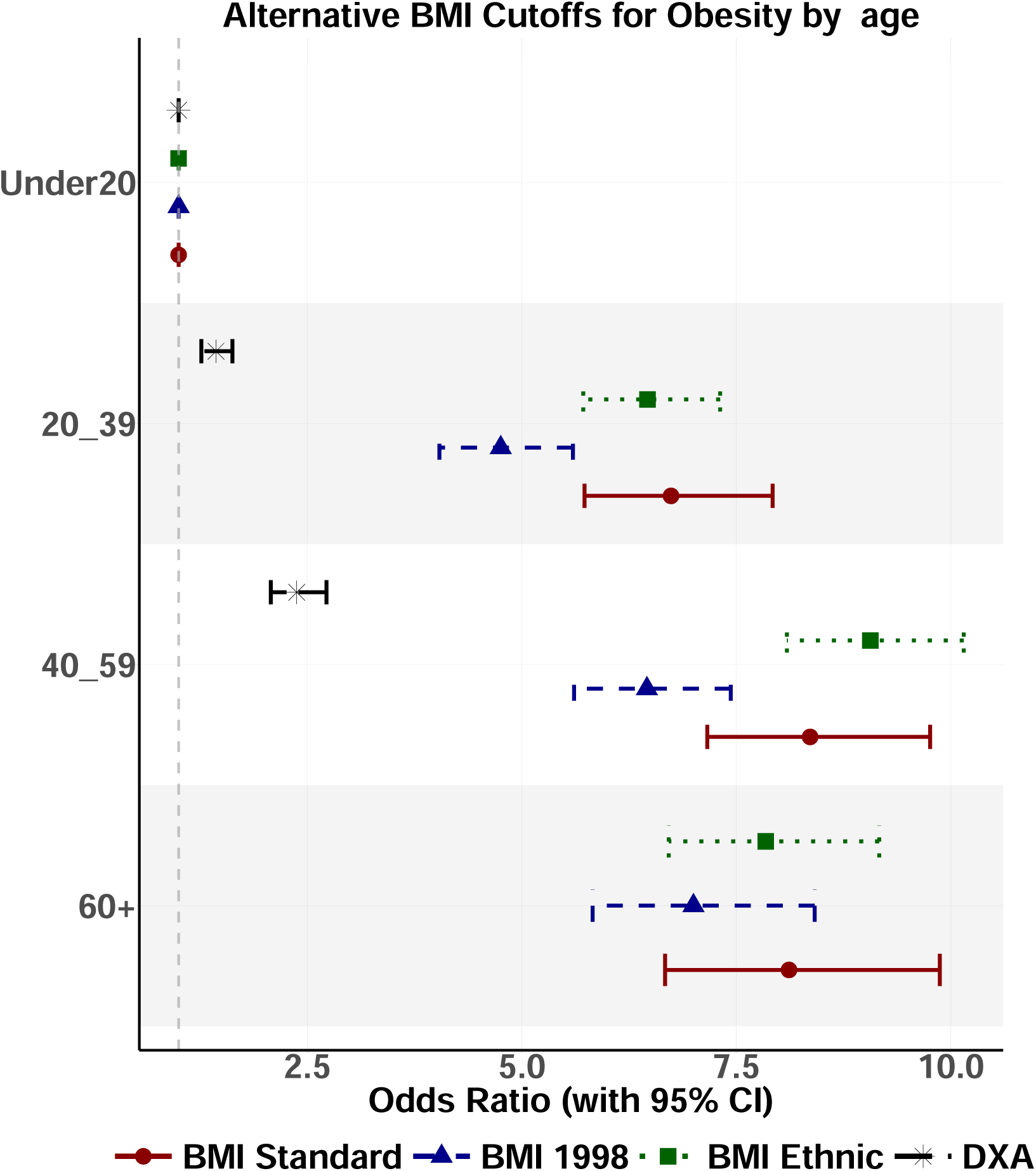
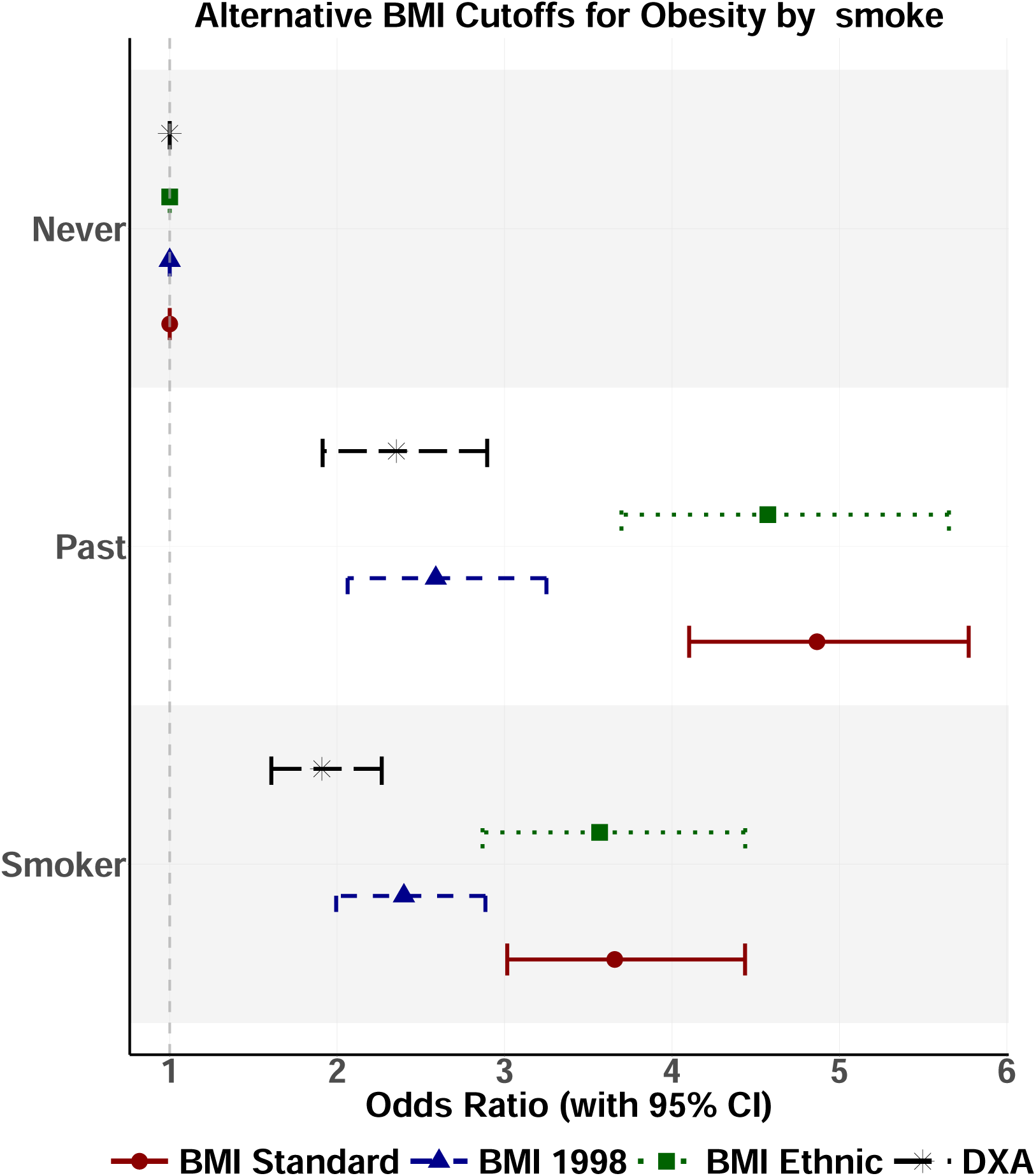
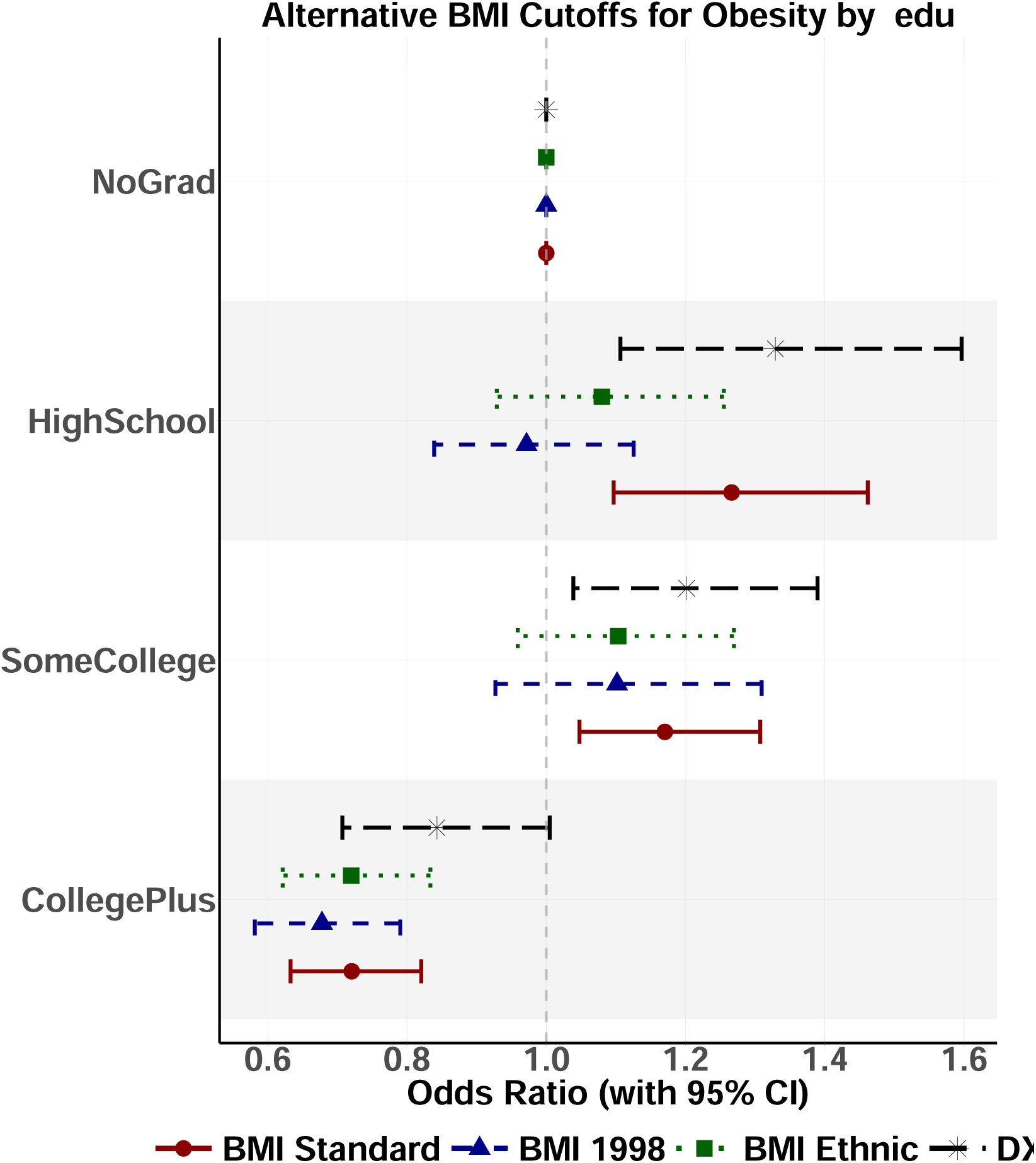
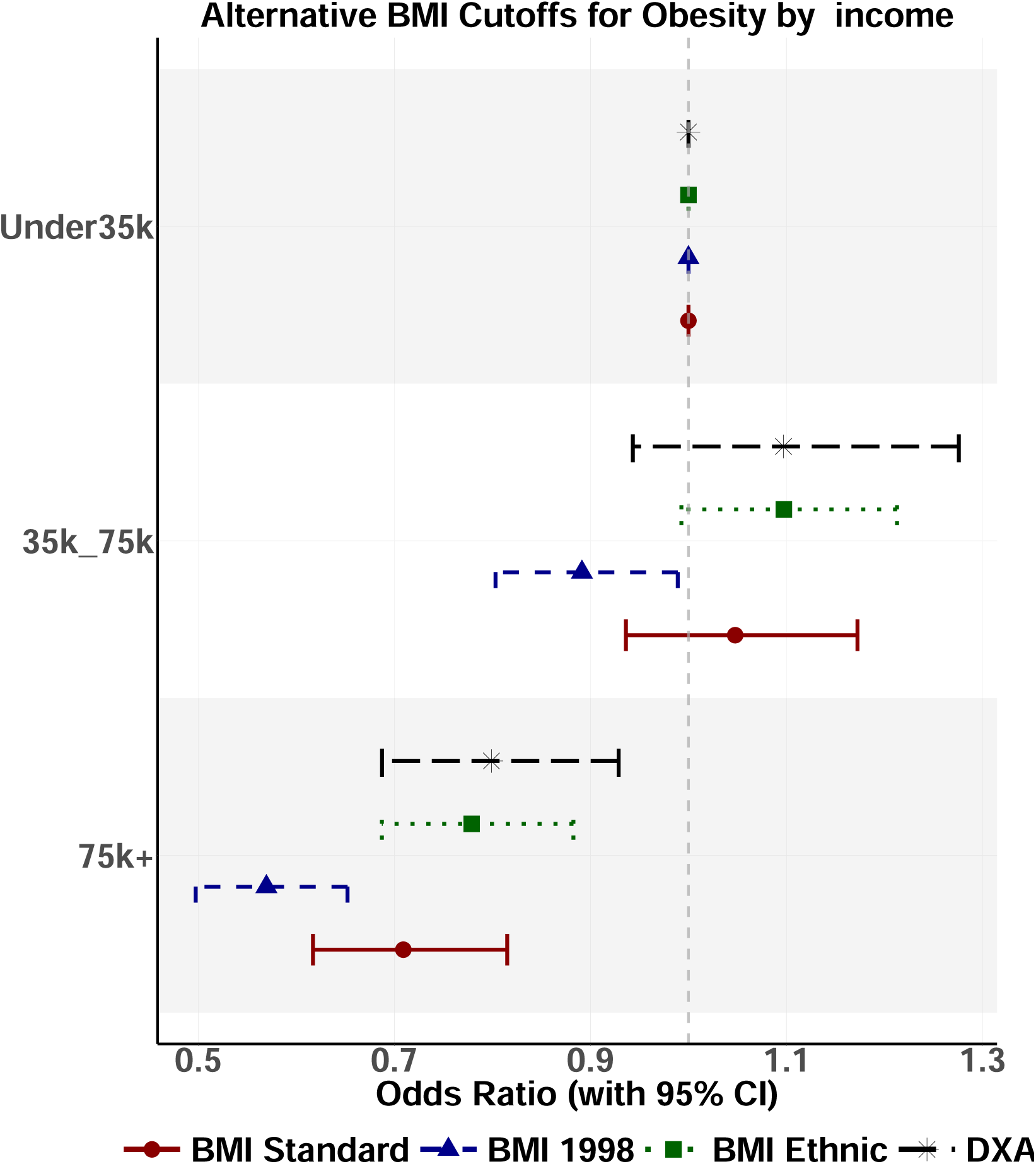

## A.7 IPD in-sample validation

Different proportions of labeled data for the 2011-2017 in-sample validation exercise are shown below (50%-50% and 30%-70% labeled-unlabeled). IPD more closely approximates DXA as the proportion of labeled data increases. As the proportion decreases, the IPD estimates become noisier and more biased, reflecting the additional uncertainty owed to the fact that the BMI are predicted data.

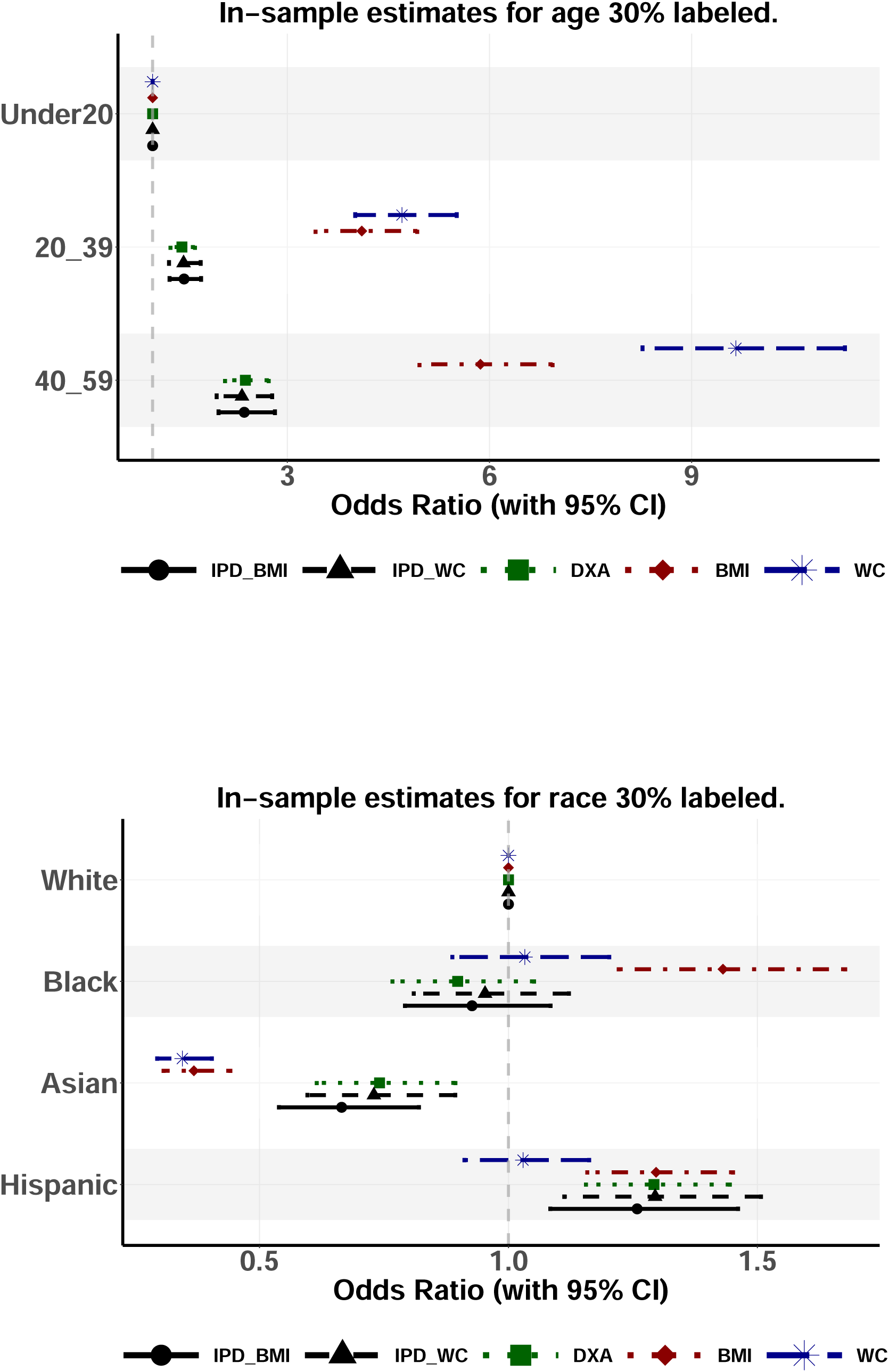
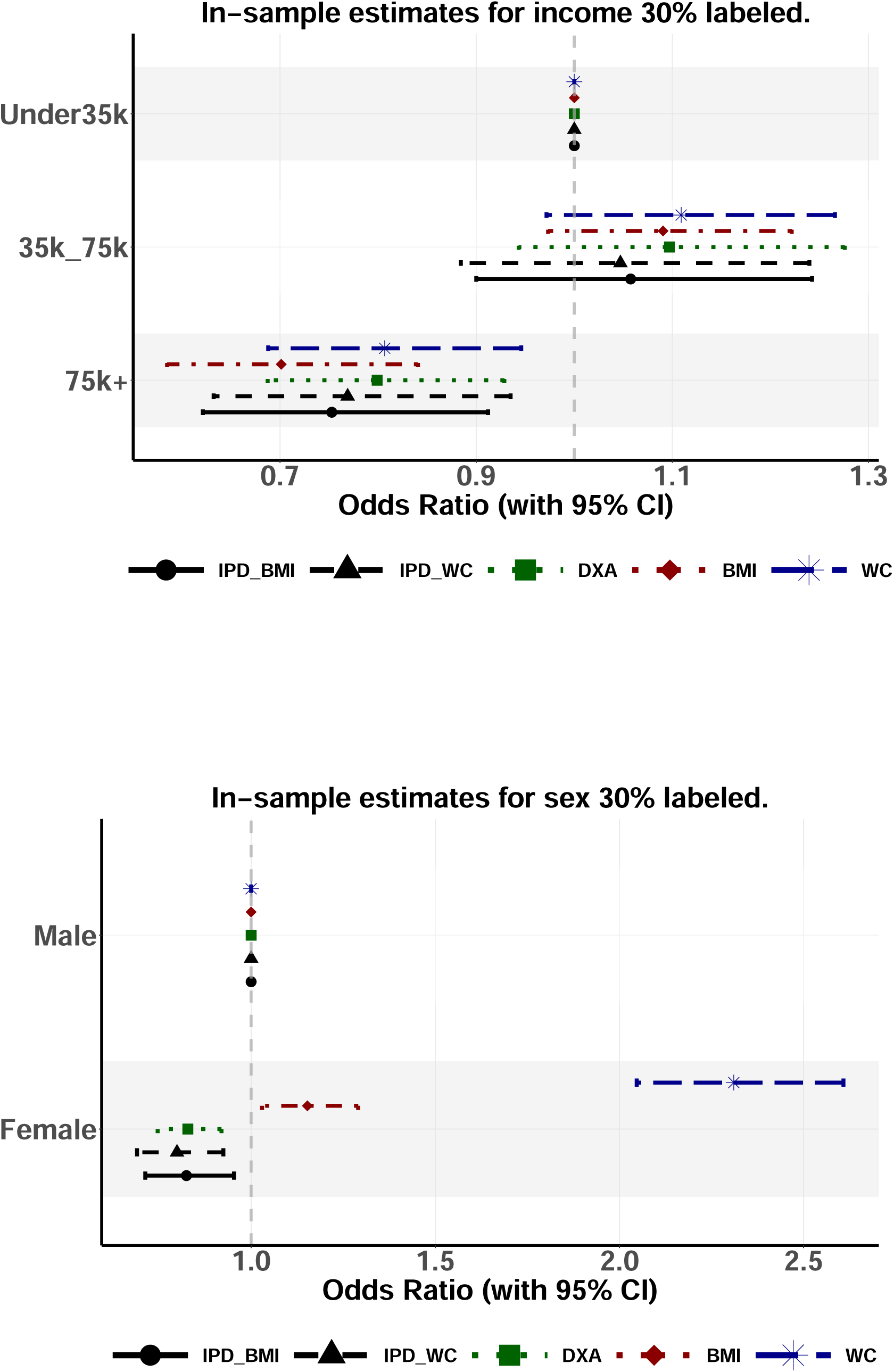
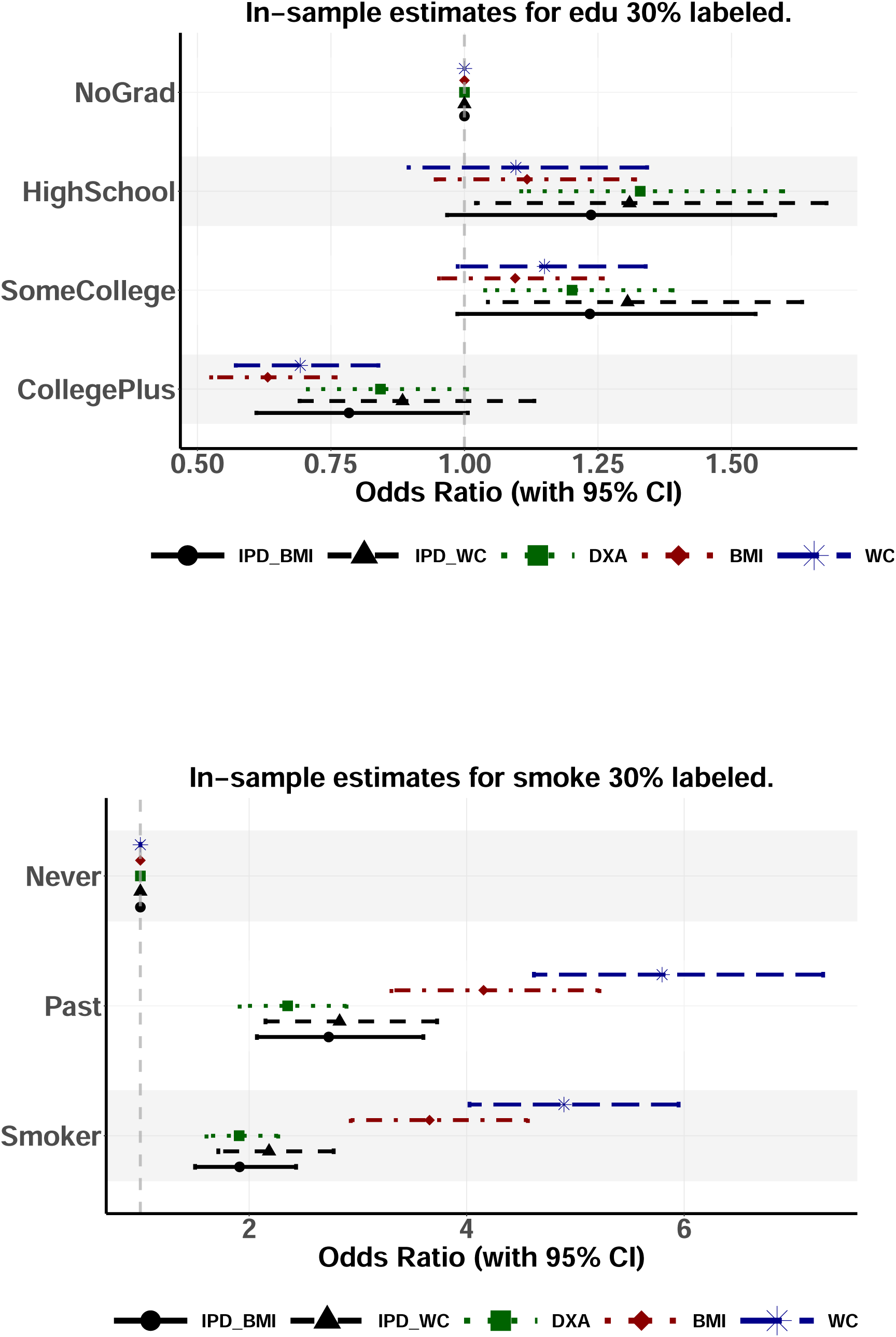
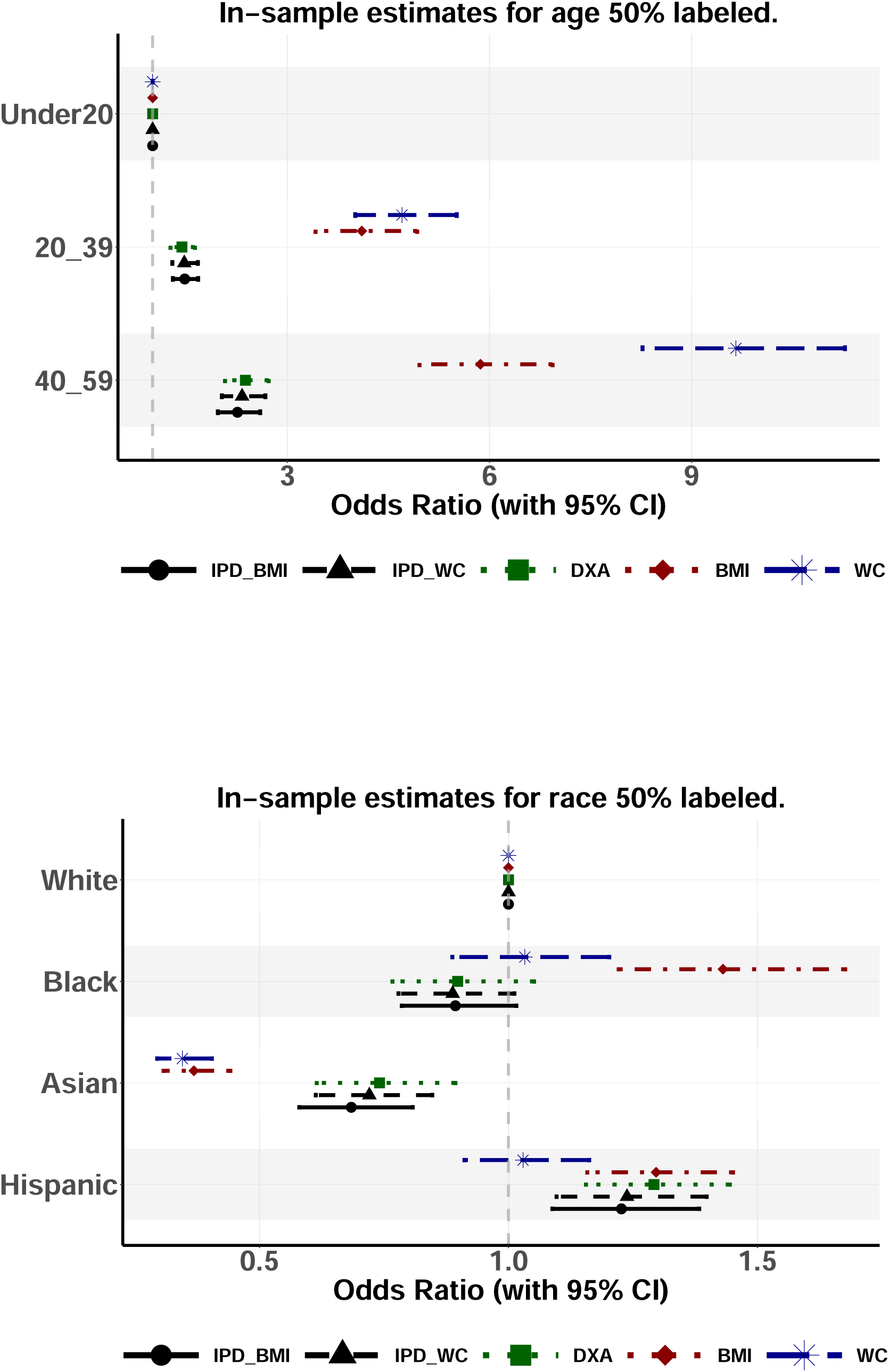
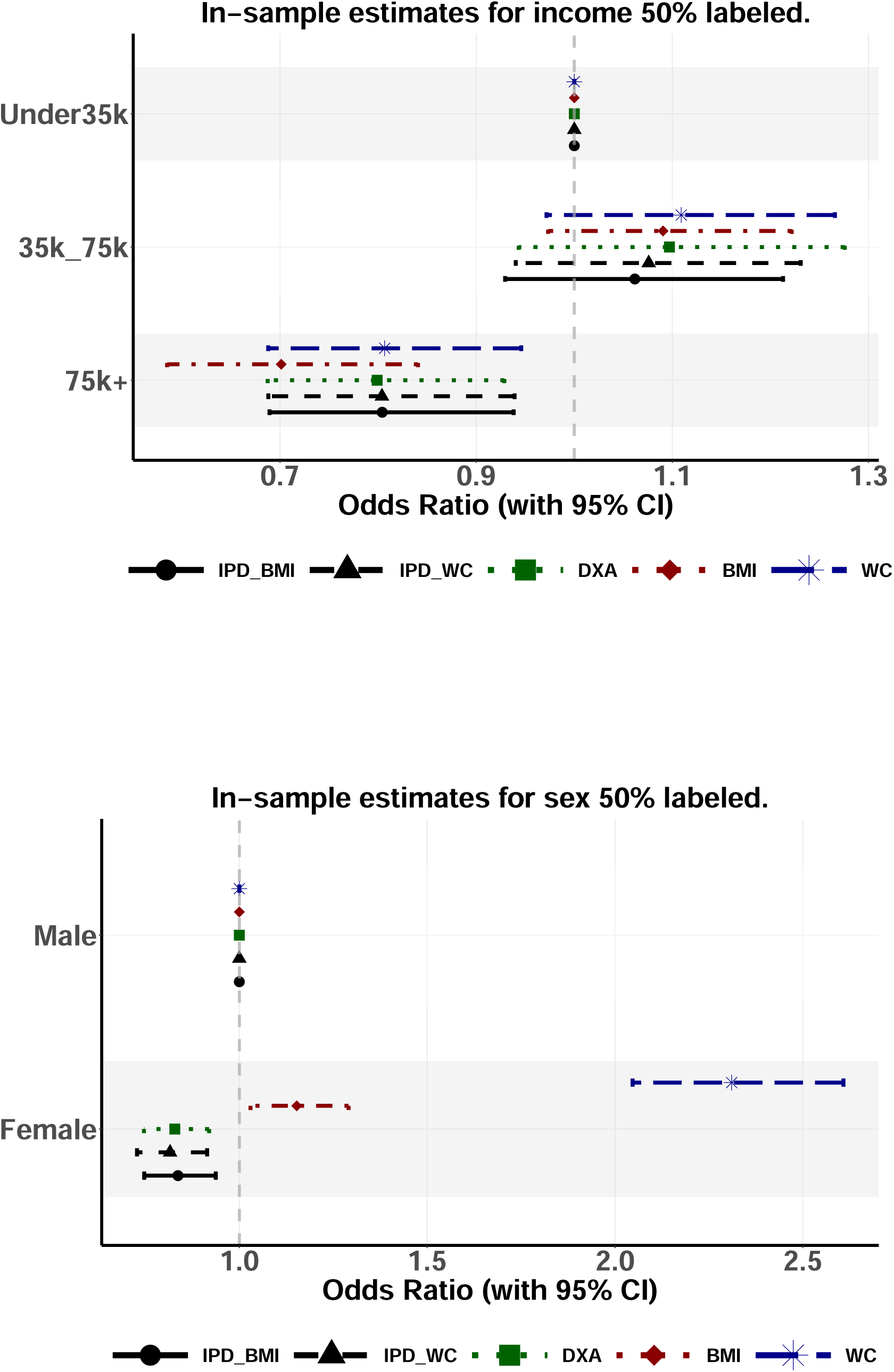
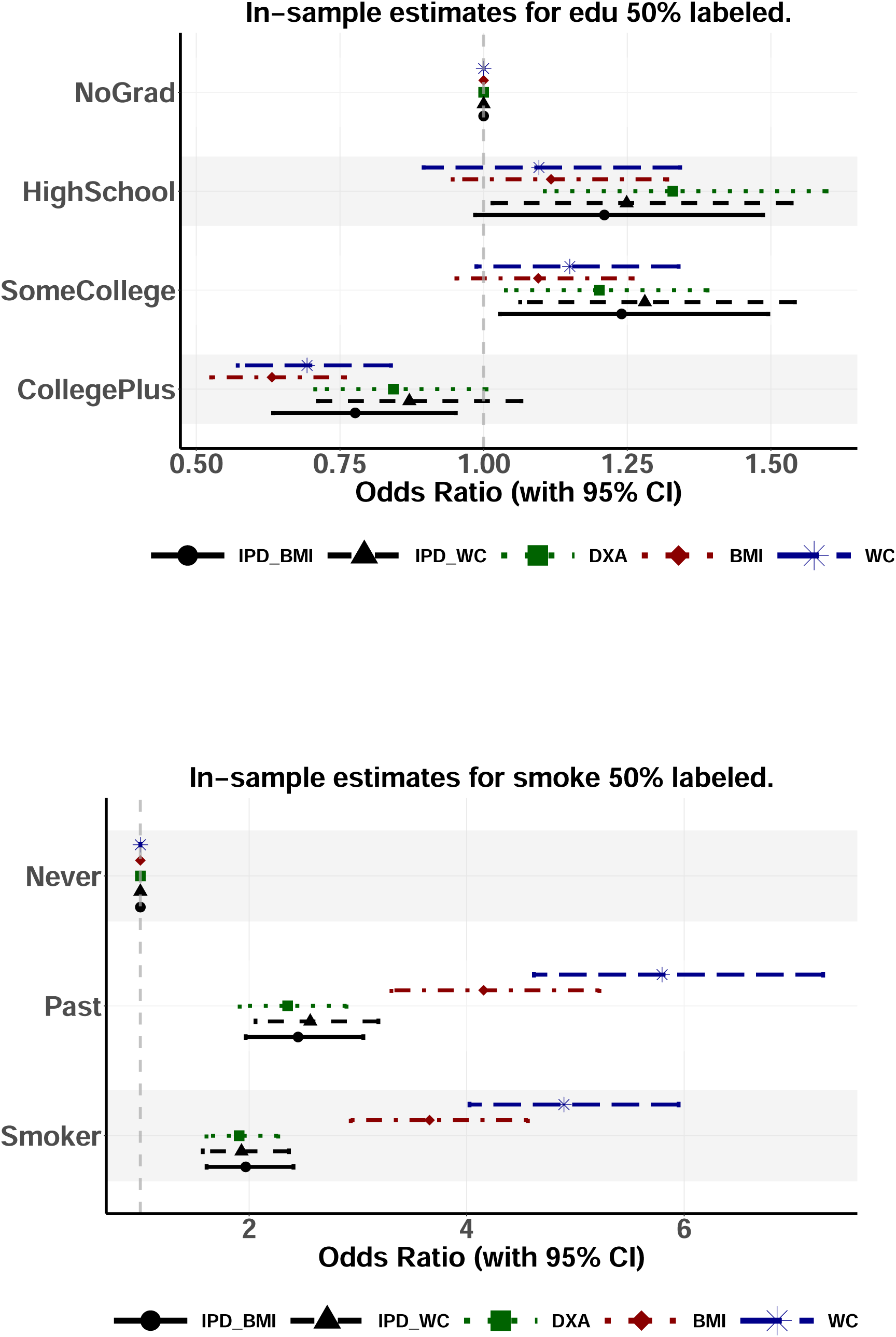

